# Trends in analgesia prescribing in primary care in Ireland and England between 2014 and 2022 - a repeated cross-sectional study

**DOI:** 10.1101/2024.06.04.24308266

**Authors:** Molly Mattsson, Michelle Flood, Brian MacKenna, Emma Wallace, Fiona Boland, Ciara Kirke, Mary E Walsh, Tom Fahey, Frank Moriarty

## Abstract

**Background:** Pain is a major public health issue, and a common reason people seek medical care. Pharmacological treatments depend on the type of pain and carry different risks and benefits. The aim of this study was to examine trends in analgesic prescribing in Ireland and England between 2014 and 2022.

**Methods:** Monthly data on medicines prescribed and dispensed in primary care were used. For Ireland, data on the means-tested General Medical Services (GMS) scheme were used, covering approximately 32% of the population. For England, data from the NHS Digital platform for all general practices were used. Outcomes were the volume of prescribed analgesic use, including rates of dispensings, costs, and standard doses (including oral morphine equivalents (OMEs) for opioids) per 1,000 population, summarised per year for each drug class/drug.

**Results:** In Ireland, the rate of analgesia dispensings increased between 2014 and 2022 for most drugs. Opioid dispensings increased from 979 to 1,220 per 1,000 population, while paracetamol increased from 1,295 to 1,824. Systemic NSAIDs decreased from 781 to 734. In England, most analgesia dispensing rates decreased, with opioids decreasing from 721 to 585 per 1,000 population, paracetamol from 734 to 484, and systemic NSAIDs from 259 to 167.

**Discussion:** Substantially different dispensing patterns were found in Ireland and England, with higher increasing overall rates in Ireland and lower decreasing rates in England, potentially driven by the older age and lower socioeconomic status of GMS patients in Ireland. Further research to understand drivers for this higher volume of use is required.

## Background

Pain is a major public health issue with a significant burden and a common reason people seek medical care(St. Sauver et al., 2013). Acute pain has a sudden onset, short duration, and is clearly associated with a cause. Chronic pain is pain that lasts or recurs for longer than three months and includes pain of uncertain origin(Treede et al., 2015). An analysis of the United States 2019 National Health Interview Survey (NHIS) data estimated that 20.5% of adults had chronic pain, with higher prevalence reported among women, older adults, and individuals with lower socioeconomic status (SES)(Yong et al., 2022), while a UK systematic review reported a pooled estimate of 43.5% for chronic pain(Fayaz et al., 2016).

Pharmaceutical treatment for pain conditions depends on the type of pain experienced. Nociceptive pain, including degenerative changes, traumatic pain, and muscle spasms, may be acute or chronic, and treated by a combination of opioids and a range of non-opioid medications, including paracetamol and topical and oral NSAIDs. In Ireland, a maximum of four days of opioids for acute pain is routinely recommended as part of a balanced multi-modal analgesia regime(National Clinical Programme for Anaesthesia, 2022). Opioids are however no longer considered to be a first-line treatment for any form of chronic pain, and many guidelines do not recommend them at all in certain populations(Rosenberg et al., 2018). Indicated treatments for neuropathic pain include certain analgesic antidepressants and antiepileptic drugs, as well as topical lidocaine and capsaicin. For nociplastic pain, e.g. fibromyalgia, non-pharmacological treatments are generally seen as preferred management, although pharmacological treatments typically used for neuropathic pain have been found to be beneficial(Bułdyś et al., 2023).

Quality of prescribing in analgesia is an important consideration. The Organisation for Economic Co-operation and Development (OECD) has outlined 11 quality indicators for prescribing in primary care, including opioid and NSAID prescribing(OECD, 2017). Opioid analgesics carry a risk of dependency and are subject to additional legislation and regulations. A systematic review has estimated rates of abuse and misuse at between 20% and 30%, and addiction at between 8% and 12%(Vowles et al., 2015)In Ireland, recent research has shown that strong opioid prescribing prevalence increased from 14.4% in 2010 to 16.3% in 2019, with the greatest increase among those ≥65 years(Norris et al., 2021). NSAIDs have been associated with severe adverse effects, in particular when used in conjunction with anticoagulants(Davis & Robson, 2016).

Prescribing of different types of analgesia can vary between countries, and may reflect inter-country differences, including practice guidelines, drug regulations, availability and timely access to non-pharmacological therapies, and resources to support discontinuation (Jayawardana et al., 2021; Ju et al., 2022; Vitiello, 2008). Cross-national comparative (CNC) studies have been commonly used to identify the variation in medication use between countries, with the aim of enhancing our understanding of the implications of varying prescribing and developing strategies to optimise medication use(Vlahovic-Palcevski et al., 2016). The aim of this study was to examine trends in analgesic prescribing in Ireland and England between 2014 and 2022.

## Methods

This was a repeated cross-sectional study of medicines dispensing in primary care in two countries, Ireland and England. The study period was 2014 to 2022. This study forms part of a larger project focused on analgesic and sedative prescribing, the protocol for which has been previous published(Mattsson, Boland, et al., 2022). The study was approved by the RCSI University of Medicine and Health Sciences Research Ethics Committee (ref: REC202201015) and the Health Service Executive (HSE) Reference Research Ethics Committee B (ref: RRECB1022FM).

### Health Service Executive data (Ireland)

Data for Ireland were obtained from the Primary Care Reimbursement Service (PCRS) for dispensing on the General Medical Services (GMS) scheme. The PCRS is the section within the health service which administers community drug schemes in Ireland, including the GMS scheme, which covers approximately 32% of the population. Eligibility for the GMS scheme is means tested, with higher thresholds for individuals aged 66 and over, and therefore eligible persons tend to be older and more socioeconomically deprived than the general population(Mattsson, Flood, et al., 2022; Sinnott et al., 2017). For adults aged 70 years and over, the scheme covers the vast majority of adults. About 10% of people eligible for the GMS scheme were eligible at a discretionary basis for individuals incurring substantial medical expenses(PCRS, 2024). Prescriptions include those initiated and prescribed by the individual’s GP, as well as those initiated by specialist services, such as pain clinics. As all prescriptions need to be transcribed by GP onto a GMS it is not possible from this data to identify who has initiated the analgesic prescribing and/or if the patient is under specialist care. Pharmacies transmit claims for prescribed medications which were dispensed to individuals eligible for community drug schemes to the PCRS at the end of each calendar month for reimbursement, and the data therefore provide complete information on prescribed medications that are dispensed to eligible people. The data contain drug information (WHO Anatomical Therapeutic Chemical (ATC) code, strength, defined daily dosage (DDD), and product information), quantity dispensed, the number of dispensings, month of dispensing, and cost. The data were not publicly available, and were provided by the PCRS following an information request made for the purposes of this project.

### National Health Service data (England)

National Health Service (NHS) data from England were extracted from the English Prescribing Dataset on the NHS Business Services Authority (NHSBSA) website, which provides monthly statistics of prescribing of different medications aggregated at the level of GP practices for all practices in England(NHS, 2022). The data relate to NHS prescriptions issued by general practices in England (by any practice prescribing staff) and dispensed in any community pharmacy in the UK. Prescribed products are coded based on their British National Formulary (BNF) classification. For this study, the items variable within the NHS Digital data, which corresponds to the number of items of each prescribed product that was dispensed in the specified month, was used, in addition to the quantity dispensed and the cost (based on net ingredient cost). A prescription item refers to a single supply of a medicine, dressing or appliance prescribed on a prescription form. The items variable does not provide any indication of the length of treatment or quantity of medicine prescribed, however often these will be issued for one month. The data were publicly available and were extracted for the relevant years and BNF codes using an R script adapted from the OpenDataAPIQuery script developed by the NHSBSA to query its open data portal API(NHSBSA, 2020).

### Outcomes

Outcomes were derived to capture volume of prescribed analgesic utilisation. Analgesics were identified using WHO ATC codes in data for Ireland and mapped to the equivalent BNF codes for England, and included opioids, systemic NSAIDs, paracetamol, topical analgesics, antimigraine preparations, and other analgesics used for neuropathic pain (see Supplementary Table 1 for the included drugs with ATC and BNF codes).

Outcomes included number of dispensings, costs, and standard doses per day, expressed as rates per 1,000 patients. For Ireland, denominators were obtained from PCRS annual reports which report number of GMS eligible individuals at the end of each calendar year(PCRS, 2023). For England, denominators were obtained from the NHS Digital portal and included all patients registered with a GP practice for each year(NHS Digital., 2023). Standard doses provide a fixed unit of measurement dependent on prescription quantity, pack size and strength to capture medications consumption on a common scale across different medications, different drugs, or different drug classes. The World Health Organisation DDD, defined as “the assumed average maintenance dose per day for a drug used for its main indication in adults”(WHO, 2018) was used for all medications, with the exception of topical analgesics. In addition, for opioids, oral morphine equivalents (OME), as per Supplementary Table 2, were used. This was applied to both single and combination medications, with strengths mapped to standard doses for each component ingredient for combinations (i.e. for paracetamol and codeine combinations, the total DDD reflects the number of DDDs of paracetamol and the number of DDDs of codeine dispensed).

### Analysis

The outcomes above were calculated in each country for each year from 2014 to 2022. Outcomes were derived for all drug classes (as presented in Supplementary Table 1). For opioids, outcomes were further calculated for each individual drug, as well as weak and strong opioids (as presented in Supplementary Table 1) and long-acting opioid formulations (slow-release oral formulations and transdermal patches). For systemic NSAIDs, outcomes were calculated for COX-2 inhibitors (i.e. coxibs) and non-selective NSAIDSs, and for topical products, for lidocaine plasters, capsaicin, and topical NSAIDs. For paracetamol, all products containing paracetamol were included, i.e. both single ingredient and combination products. Individual indicators were derived for other analgesics often used in neuropathic pain, including pregabalin, gabapentin, and amitriptyline 10mg. Typically, higher doses of amitriptyline are used to treat depression, whereas low doses are prescribed for chronic pain(McQuay et al., 1996). Although doses higher than 10mg may be prescribed for pain, we have limited analysis to 10mg as this dose is not used for depression treatment, thus ensuring that no prescriptions not prescribed for pain management are included.

Rates for each outcome were summarised per year for each drug class and drug in both countries. The absolute and relative change from 2014 to 2022 was calculated for each, as well as the rate ratio (the rate in Ireland divided by the rate in England). In addition, the percentage share of analgesic prescribing accounted for by each drug class and drug was calculated per year, as well as the percentage share of opioid prescribing accounted for by each opioid drug. The Jonckheere–Terpstra test for linear trend across years was run for each drug/drug class separately for Ireland and England. Considering data from Ireland relates to the GMS scheme, which overrepresents those with lower SES, a subgroup analysis was conducted using data for England from higher deprivation clinical commissioning groups (CCGs) only (those CCGs in the top 33 centiles of deprivation, based on the Index of Multiple of Deprivation 2019(UK Goverment, 2019).

## Results

In Ireland, the number of GMS eligible patients during the study period decreased from 1,768,700 in 2014 to 1,568,379 in 2022. Conversely, the NHS population in England increased year-on-year from 56,545,892 in 2014 to 61,768,942 in 2022. See Supplementary Table 3 for populations for both countries between 2014 and 2022.

In Ireland, the rate of analgesia dispensings per 1,000 GMS population increased from 3,388.6 in 2014 to 4,316.0 in 2022. Opioid dispensings increased from 978.7 per 1,000 population in 2014 to 1220.2 in 2022. Dispensing of most individual opioid increased, with the exception of hydromorphone (3.4 per 1,000 in 2014 vs 1.5 in 2022), dihydrocodeine (40.5 per 1,000 in 2014 vs 0.01 in 2022), and tramadol (271.3 per 1,000 in 2014 vs 256.7 in 2022). Topical analgesics, low-dose amitriptyline, gabapentin, pregabalin, and paracetamol similarly all increased, while systemic NSAIDs decreased (781 per 1,000 in 2014 vs 733.7 in 2022). Conversely, in England, the rate of analgesia dispensings per 1,000 population decreased from 1,834.0 in 2014 to 1,516.5 2022, opioid dispensing decreased from 720.5 to 584.6, with topical analgesics, paracetamol, and systemic NSAIDs all decreasing as well. Most opioids also decreased, while very small increases were seen for morphine, oxycodone, buprenorphine, and tapentadol. Low-dose amitriptyline, gabapentin, and pregabalin saw increased dispensings throughout the study period (123.5 per 1,000 in 2014 vs 166 in 2022, 87 vs 109.5, and 71.3 vs 126.7 respectively).

In Ireland, the largest relative increases between 2014 and 2022 were seen in dispensing of tapentadol (389%), 10 mg amitriptyline (194%), gabapentin (97%), topical NSAIDs (59%), and oxycodone (50%). The largest relative decreases in dispensings were seen for dihydrocodeine (99%), lidocaine plasters (83%), and hydromorphone (54%). In England, tapentadol (130%) and pregabalin (78%) increased the most, while pethidine had the largest decrease (80%). See Figures 1a and 1b and 2a and 2b and Tables 1a and 1b for dispensings for both countries for all years included in the study. Further, we calculated absolute and relative differences for absolute dispensings for both Ireland and England. For Ireland, absolute dispensings increased to a lesser degree than the dispensing rates. Conversely, absolute dispensings decreased to a lesser extent compared to the dispensing rates in England. See Supplementary Table 4.

**Figure 1a.**
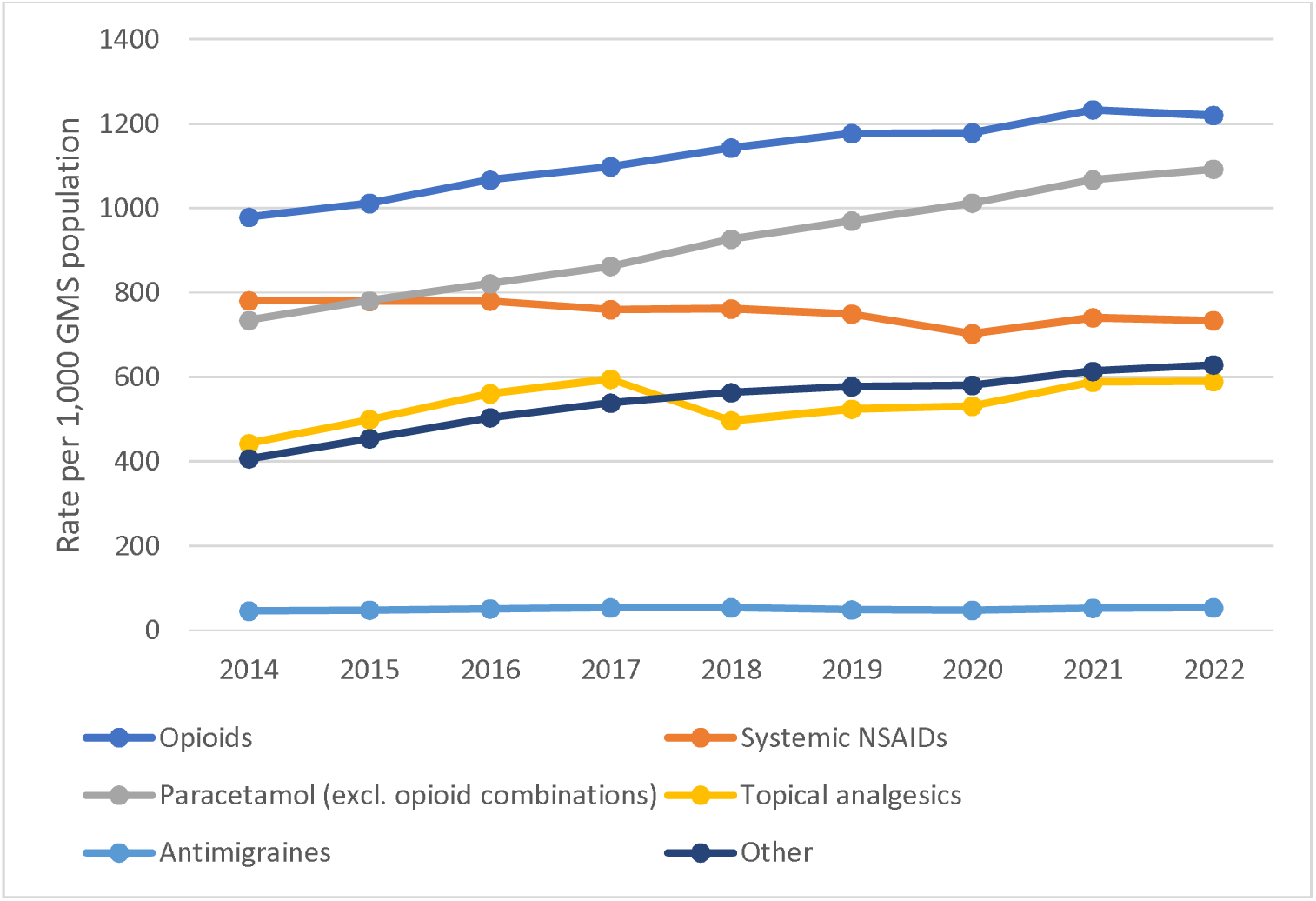
Analgesic dispensings 2014-2022 (Ireland)

**Figure 1b.**
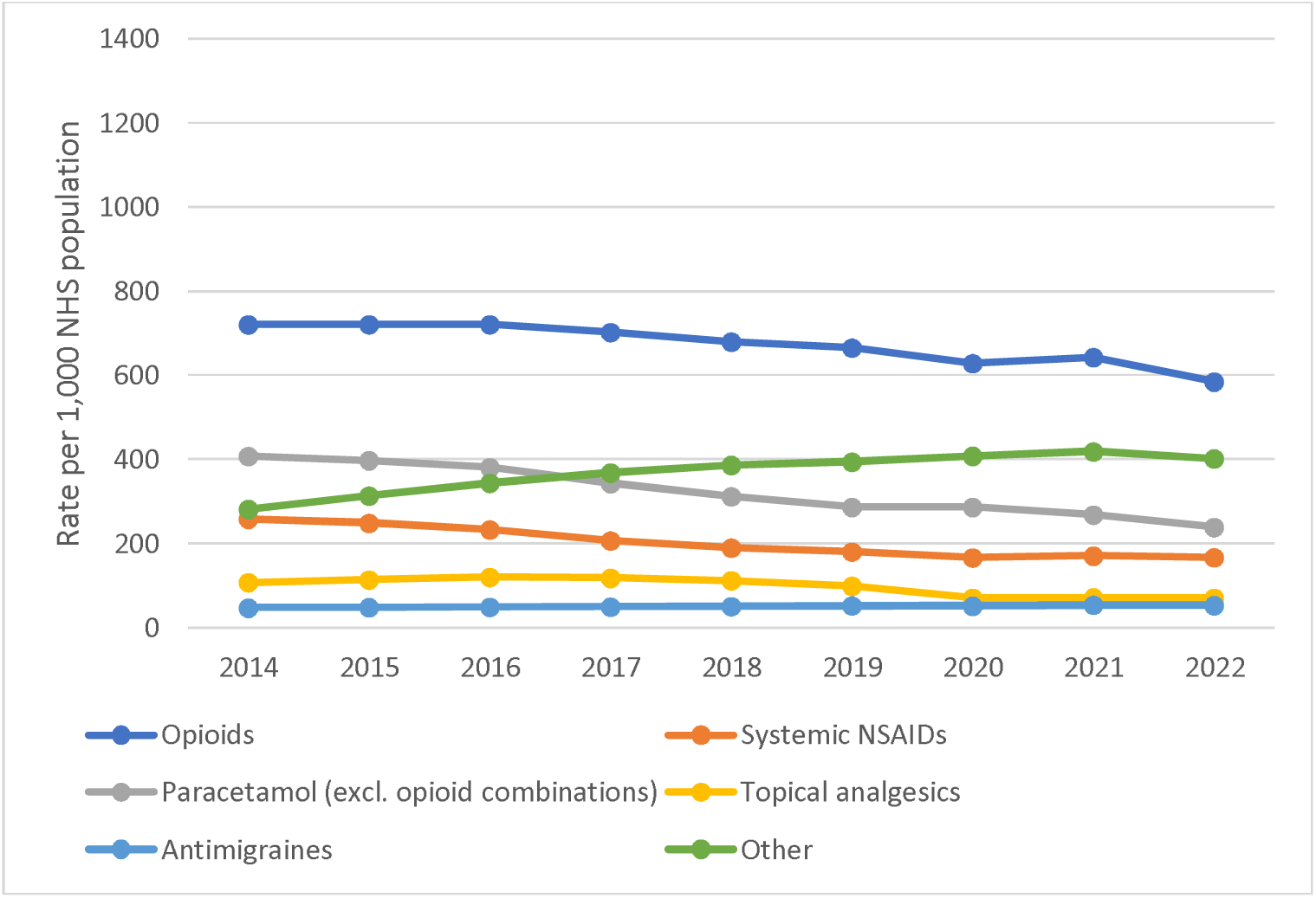
Analgesic dispensings 2014-2022 (England)

**Figure 2a.**
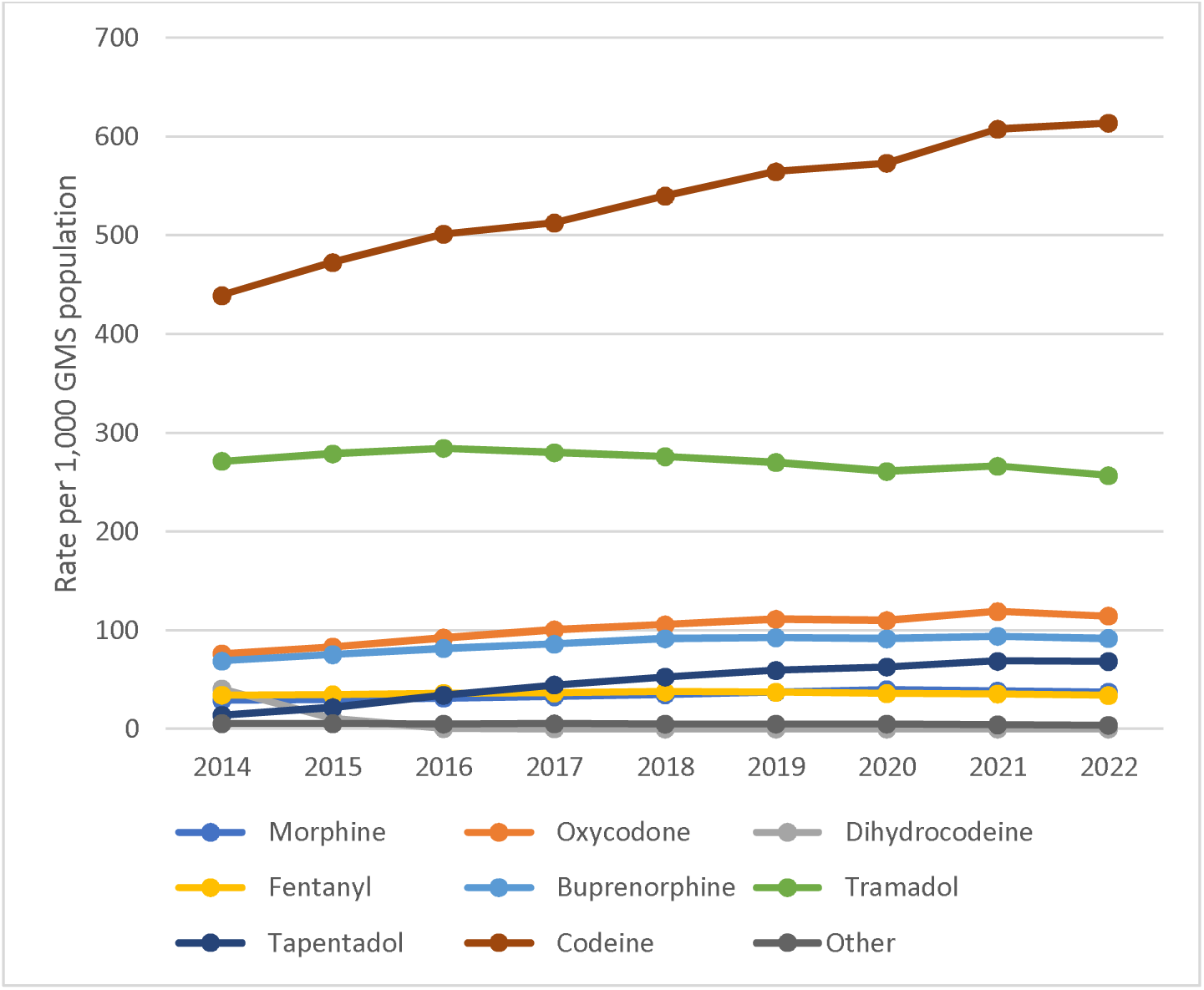
Opioid dispensings 2014-2022 (Ireland)

**Figure 2b.**
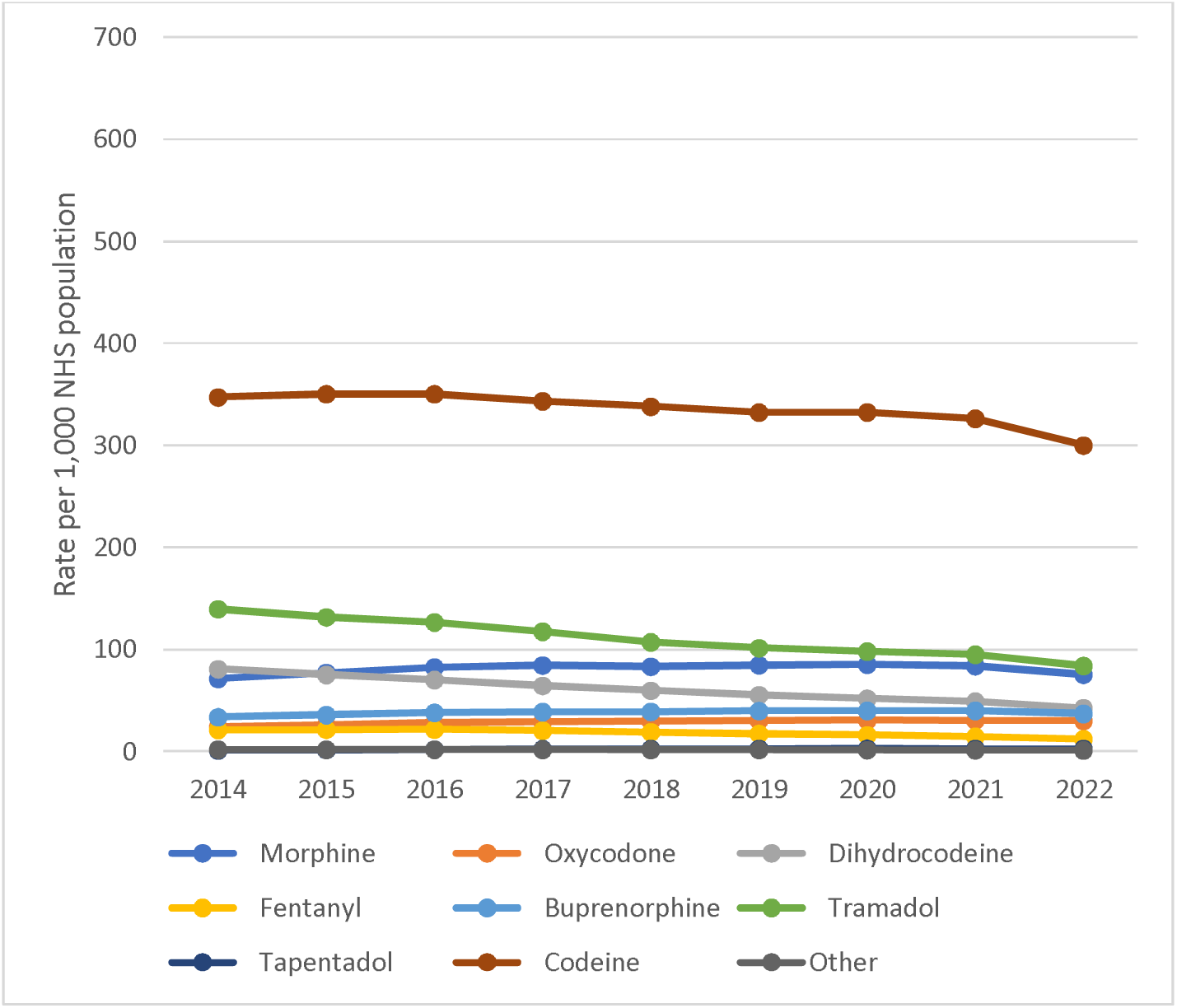
Opioid dispensings 2014-2022 (England)

**Table 1a.** Rate of dispensings per 1,000 GMS population in Ireland.

|  | 2014 | 2015 | 2016 | 2017 | 2018 | 2019 | 2020 | 2021 | 2022 | Absolute change | Relative change |
| --- | --- | --- | --- | --- | --- | --- | --- | --- | --- | --- | --- |
| All analgesics | 3388.64 | 3573.87 | 3783.35 | 3907.13 | 3944.40 | 4044.44 | 4051.22 | 4293.03 | 4316.04 | 927.40* | 0.27 |
| Opioids | 978.65 | 1011.90 | 1066.79 | 1098.57 | 1142.95 | 1177.24 | 1178.98 | 1233.47 | 1220.16 | 241.51* | 0.25 |
| *Strong opioids<sup>†</sup>* | 497.13 | 526.82 | 562.90 | 583.22 | 600.50 | 610.11 | 603.28 | 623.49 | 604.05 | 106.91* | 0.22 |
| *Long-acting opioids* | 203.19 | 217.61 | 237.45 | 250.97 | 263.36 | 268.90 | 265.28 | 272.01 | 264.30 | 61.12* | 0.30 |
| *Morphine* | 29.01 | 29.74 | 31.32 | 32.85 | 34.70 | 37.20 | 39.72 | 38.37 | 37.27 | 8.27* | 0.28 |
| *Hydromorphone* | 3.37 | 3.23 | 2.84 | 2.58 | 2.41 | 2.40 | 2.52 | 1.97 | 1.54 | -1.83* | -0.54 |
| *Oxycodone* | 76.23 | 83.11 | 92.22 | 100.53 | 105.55 | 111.22 | 109.96 | 118.97 | 114.43 | 38.20* | 0.50 |
| *Dihydrocodeine* | 40.47 | 10.57 | 0.55 | 0.10 | 0.05 | 0.03 | 0.01 | 0.01 | 0.01 | -40.46* | -1.00 |
| *Pethidine* | 0.14 | 0.18 | 0.19 | 0.13 | 0.12 | 0.04 | 0 | 0 | 0 |  |  |
| *Fentanyl* | 34.37 | 34.63 | 36.18 | 36.65 | 37.52 | 37.33 | 35.92 | 35.49 | 33.94 | -0.43 | -0.01 |
| *Buprenorphine* | 68.74 | 75.45 | 81.58 | 85.89 | 91.48 | 92.55 | 91.38 | 93.95 | 91.75 | 23.02* | 0.33 |
| *Tramadol* | 271.29 | 278.76 | 284.36 | 279.99 | 275.99 | 270.07 | 260.89 | 266.12 | 256.66 | -14.63* | -0.05 |
| *Meptazinol* | 2.00 | 2.02 | 2.07 | 2.61 | 2.46 | 2.67 | 2.53 | 2.33 | 2.28 | 0.28 | 0.14 |
| *Tapentadol* | 13.99 | 21.72 | 34.21 | 44.59 | 52.72 | 59.31 | 62.89 | 68.62 | 68.45 | 54.46* | 3.89 |
| *Codeine* | 439.05 | 472.49 | 501.27 | 512.63 | 539.95 | 564.44 | 573.15 | 607.64 | 613.83 | 174.78* | 0.40 |
| Systemic NSAIDs | 781.02 | 779.95 | 780.41 | 760.08 | 761.69 | 749.53 | 702.89 | 740.69 | 733.67 | -47.35* | -0.06 |
| *Non-selective NSAIDs* | 665.63 | 661.13 | 659.78 | 637.88 | 639.69 | 630.07 | 584.43 | 618.44 | 615.07 | -50.57* | -0.08 |
| *Coxib* | 115.39 | 118.82 | 120.63 | 122.20 | 122.00 | 119.46 | 118.46 | 122.25 | 118.60 | 3.21 | 0.03 |
| Paracetamol (incl. combinations) | 1294.57 | 1351.98 | 1421.31 | 1477.08 | 1570.05 | 1638.32 | 1691.39 | 1789.34 | 1823.81 | 529.24* | 0.41 |
| *Paracetamol (excl. opioid combinations)* | 734.34 | 780.61 | 821.14 | 862.32 | 926.88 | 969.85 | 1012.52 | 1067.24 | 1092.78 | 358.44* | 0.49 |
| Gabapentin | 54.58 | 58.20 | 64.24 | 71.46 | 78.51 | 84.96 | 91.20 | 101.71 | 107.71 | 53.13* | 0.97 |
| Pregabalin | 309.31 | 344.34 | 378.04 | 393.54 | 400.39 | 396.80 | 385.30 | 397.83 | 395.62 | 86.31* | 0.28 |
| Low-dose amitriptyline | 42.67 | 51.55 | 61.45 | 73.21 | 84.25 | 95.47 | 104.04 | 114.10 | 125.67 | 82.99* | 1.94 |
| Topical analgesics | 442.41 | 499.41 | 560.66 | 594.93 | 496.42 | 524.07 | 531.08 | 587.91 | 589.38 | 146.97 | 0.33 |
| *Topical NSAIDs* | 357.32 | 381.12 | 405.74 | 440.35 | 474.79 | 501.22 | 512.13 | 566.34 | 568.45 | 211.13* | 0.59 |
| *Lidocaine* | 78.56 | 111.46 | 147.87 | 146.20 | 10.43 | 14.17 | 14.56 | 13.90 | 13.02 | -65.55 | -0.83 |
| *Capsaicin* | 6.52 | 6.83 | 7.05 | 8.39 | 11.19 | 8.68 | 4.38 | 7.67 | 7.91 | 1.39 | 0.21 |
| Antimigraines | 45.65 | 47.91 | 50.61 | 53.02 | 53.38 | 48.32 | 47.26 | 52.14 | 53.33 | 7.68 | 0.17 |

\*Test for linear trend significant at $p < 0.05$
<sup>†</sup>Morphine, hydromorphone, oxycodone, pethidine, fentanyl, buprenorphine, tramadol, tapentadol

**Table 1b.** Rate of dispensings per 1,000 NHS population in England.

|  | 2014 | 2015 | 2016 | 2017 | 2018 | 2019 | 2020 | 2021 | 2022 | Absolute change | Relative change |
| --- | --- | --- | --- | --- | --- | --- | --- | --- | --- | --- | --- |
| All analgesics | 1824.02 | 1843.44 | 1848.32 | 1788.89 | 1731.77 | 1678.68 | 1642.55 | 1626.29 | 1516.54 | -307.48* | -0.17 |
| Opioids | 720.51 | 720.69 | 720.60 | 702.16 | 679.70 | 665.44 | 628.37 | 642.72 | 584.59 | -135.92* | -0.19 |
| *Strong opioids<sup>†</sup>* | 291.53 | 294.02 | 298.91 | 293.25 | 281.02 | 276.73 | 242.90 | 266.43 | 241.23 | -50.29* | -0.17 |
| *Long-acting opioids* | 131.80 | 137.13 | 141.62 | 141.33 | 137.70 | 137.01 | 116.97 | 130.40 | 118.12 | -13.68 | -0.10 |
| *Morphine* | 71.30 | 77.07 | 82.36 | 84.43 | 83.11 | 84.65 | 85.34 | 83.73 | 75.33 | 4.03 | 0.06 |
| *Hydromorphone* | 0.17 | 0.17 | 0.16 | 0.17 | 0.15 | 0.14 | 0.13 | 0.11 | 0.10 | -0.07* | -0.40 |
| *Oxycodone* | 23.90 | 25.82 | 27.75 | 29.19 | 29.92 | 30.54 | 30.67 | 30.43 | 30.12 | 6.22* | 0.26 |
| *Dihydrocodeine* | 80.73 | 75.15 | 70.18 | 64.45 | 59.63 | 55.30 | 51.90 | 48.95 | 42.49 | -38.23* | -0.47 |
| *Pethidine* | 0.59 | 0.55 | 0.49 | 0.43 | 0.39 | 0.30 | 0.19 | 0.13 | 0.12 | -0.47* | -0.80 |
| *Fentanyl* | 21.26 | 21.45 | 21.52 | 20.44 | 18.85 | 17.35 | 15.94 | 14.42 | 12.08 | -9.19* | -0.43 |
| *Buprenorphine* | 33.62 | 35.82 | 38.01 | 38.75 | 38.97 | 39.73 | 39.90 | 39.96 | 36.96 | 3.34* | 0.10 |
| *Tramadol* | 139.54 | 131.63 | 126.63 | 117.54 | 107.11 | 101.32 | 98.06 | 94.96 | 83.90 | -55.64* | -0.40 |
| *Meptazinol* | 1.17 | 1.19 | 1.20 | 1.18 | 1.15 | 1.15 | 1.12 | 1.08 | 0.94 | -0.23* | -0.20 |
| *Tapentadol* | 1.14 | 1.52 | 1.98 | 2.31 | 2.51 | 2.69 | 2.74 | 2.69 | 2.63 | 1.49* | 1.30 |
| *Codeine* | 347.13 | 350.37 | 350.34 | 343.31 | 337.92 | 332.29 | 332.46 | 326.28 | 299.95 | -47.19* | -0.14 |
| Systemic NSAIDs | 258.76 | 248.90 | 233.35 | 206.93 | 190.27 | 180.65 | 166.63 | 170.65 | 167.08 | -91.68* | -0.35 |
| *Non-selective NSAIDs* | 247.08 | 237.65 | 222.17 | 196.14 | 179.45 | 169.10 | 154.58 | 157.97 | 153.83 | -93.25* | -0.38 |
| *Coxib* | 11.68 | 11.25 | 11.18 | 10.79 | 10.81 | 11.55 | 12.05 | 12.68 | 13.25 | 1.57 | 0.13 |
| Paracetamol (incl. combinations) | 733.56 | 717.82 | 693.37 | 642.79 | 601.67 | 566.28 | 564.10 | 537.55 | 484.34 | -249.21* | -0.34 |
| *Paracetamol (excl. opioid combinations)* | 407.80 | 397.71 | 380.93 | 343.29 | 312.21 | 286.55 | 286.69 | 268.43 | 238.92 | -168.88* | -0.41 |
| Gabapentin | 87.01 | 99.74 | 111.44 | 120.75 | 124.15 | 120.92 | 122.26 | 120.45 | 109.48 | 22.47 | 0.26 |
| Pregabalin | 71.30 | 83.21 | 95.15 | 106.11 | 117.58 | 123.00 | 129.46 | 135.25 | 126.67 | 55.37* | 0.78 |
| Low-dose amitriptyline | 123.53 | 130.66 | 137.40 | 141.44 | 144.84 | 150.20 | 156.44 | 163.03 | 166.00 | 42.47* | 0.34 |
| Topical analgesics | 107.92 | 114.01 | 120.16 | 118.36 | 112.40 | 99.32 | 70.97 | 71.59 | 70.64 | -37.27* | -0.35 |
| *Topical NSAIDs* | 100.22 | 105.71 | 110.75 | 108.95 | 103.83 | 93.23 | 65.43 | 64.86 | 64.69 | -35.53* | -0.35 |
| *Lidocaine* | 3.36 | 3.92 | 4.41 | 4.29 | 3.36 | 2.65 | 2.15 | 2.01 | 1.96 | -1.40* | -0.42 |
| *Capsaicin* | 4.34 | 4.39 | 5.00 | 5.12 | 5.21 | 3.44 | 3.39 | 4.71 | 3.99 | -0.34 | -0.08 |
| Antimigraines | 47.19 | 48.52 | 49.29 | 49.85 | 50.63 | 52.59 | 51.68 | 54.17 | 53.16 | 5.97* | 0.13 |

\*Test for linear trend significant at $p < 0.05$
†Morphine, hydromorphone, oxycodone, pethidine, fentanyl, buprenorphine, tramadol, tapentadol

There were large discrepancies in dispensing rates between the two populations (see Figure 2 and Supplementary Table 3). In 2022, the dispensing rate of tapentadol dispensing was 26 times higher in the GMS population in Ireland compared to England, while hydromorphone dispensing was 15 times higher. All other drugs also had higher dispensing rates in Ireland in 2022, with the exception of dihydrocodeine (rate ratio <0.01), morphine (0.49), and low-dose amitriptyline (0.76). Rate ratio was not calculated for pethidine as it was not dispensed in Ireland between 2020 and 2022. See Figure 3.

**Figure 3.**
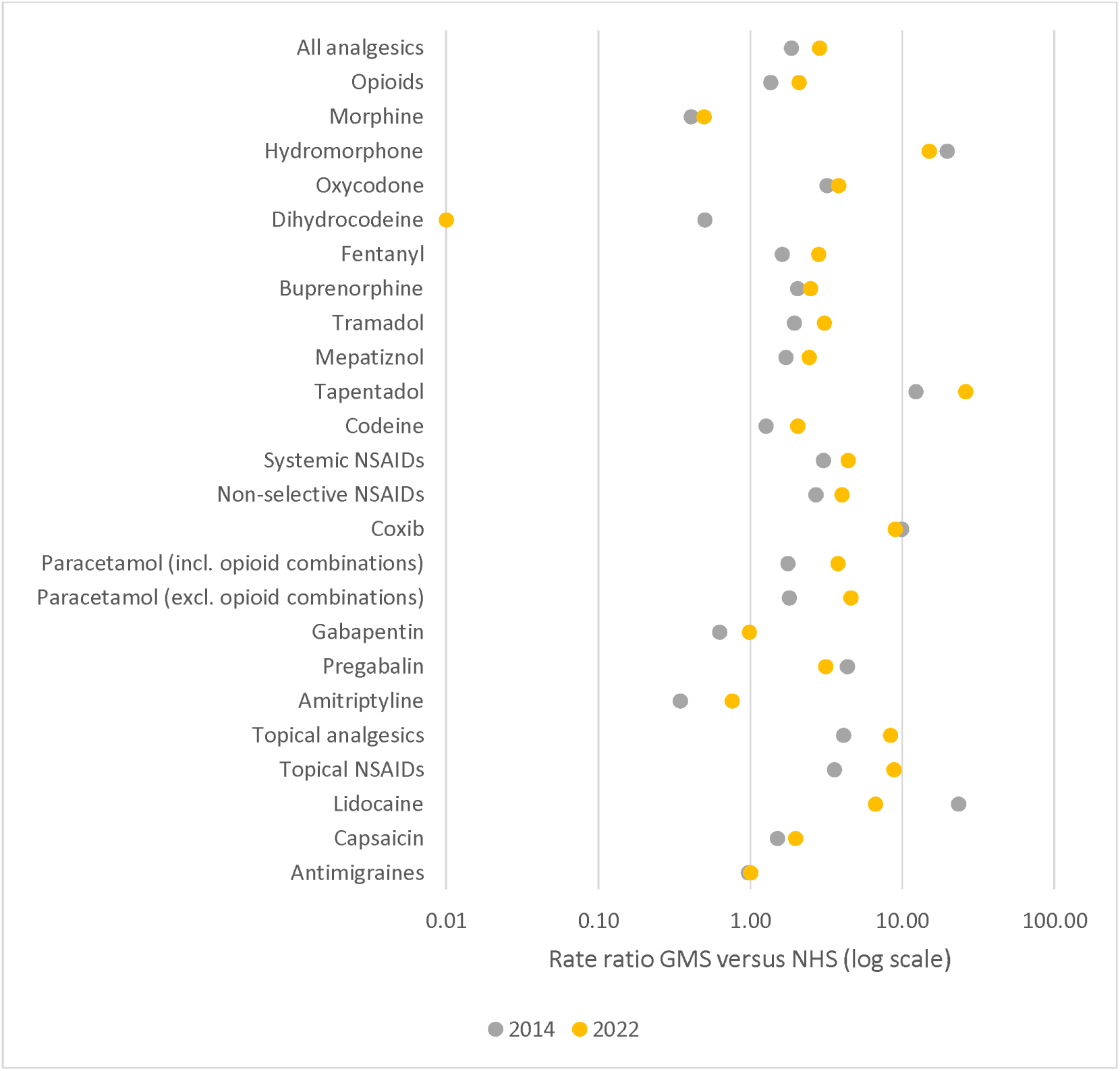
Rate ratio for dispensings in Ireland versus England, shown for 2014 (grey)and 2022 (yellow)* *Rates above 1.0 indicate higher dispensings in Ireland

The percentage share of analgesic dispensing for drug groups and individual drugs remained largely steady in both populations throughout the study period, with some exceptions. In Ireland, paracetamol dispensing accounted for 43% of analgesic utilisation in 2014, compared to 48% in 2022. Systemic NSAIDs made up 26% of analgesic dispensing in 2014, decreasing to 19% in 2022. In England, paracetamol dispensing made up 40% of analgesic utilisation in 2014, decreasing to 32% in 2022. Additionally, there were some changes in the percentage share of the individual drugs that made up opioid dispensing for both populations. In Ireland, dihydrocodeine accounted for 4% of opioid dispensing in 2014, compared to less than 1% in 2022, while tramadol decreased from 28% to 21%. Conversely, tapentadol increased from 1% to 6% and codeine from 45% to 50%. In England, tramadol dispensing decreased from 19% to 14%. See Supplementary Table 5 and Supplementary Figure 1.

In Ireland, the rate of DDD of analgesics dispensed per 1,000 GMS population per day increased from 116 in 2014 to 147.6 in 2022. Opioid DDDs increased from 33 per 1,000 population per day to 40.3. The rate of most DDDs Increased for most individual drugs, with the largest relative increases seen for tapentadol and low-dose amitriptyline. In England, the opposite was seen with DDDs per 1,000 per day decreasing from 97 to 80.6, and opioids from 39.1 to 31.9. Nearly all individual drugs decreased, while tapentadol, low-dose amitriptyline, pregabalin, and gabapentin saw increases. See Table 2a and 2b. Rate ratios for DDDs are available in Supplementary Table 6 and Supplementary Figure 2, and were similar but generally smaller in magnitude than the rate ratios for dispensing.

**Table 2a.** Rate of daily defined doses (DDDs) per 1,000 GMS population per day in Ireland.

|  | 2014 | 2015 | 2016 | 2017 | 2018 | 2019 | 2020 | 2021 | 2022 | Absolute change | Relative change |
| --- | --- | --- | --- | --- | --- | --- | --- | --- | --- | --- | --- |
| All analgesics | 130.83 | 138.61 | 145.12 | 149.73 | 155.37 | 158.30 | 159.97 | 167.75 | 166.62 | 35.78* | 0.27 |
| Opioids | 33.02 | 34.43 | 36.01 | 36.90 | 38.22 | 39.26 | 39.57 | 41.10 | 40.29 | 7.27* | 0.22 |
| *Strong opioids<sup>†</sup>* | 15.05 | 15.62 | 16.48 | 16.90 | 17.15 | 17.20 | 16.92 | 17.17 | 16.41 | 1.36 | 0.09 |
| *Long-acting opioids* | 6.59 | 6.92 | 7.37 | 7.65 | 7.86 | 7.96 | 7.82 | 7.86 | 7.59 | 1.00 | 0.15 |
| *Morphine* | 0.52 | 0.53 | 0.55 | 0.55 | 0.56 | 0.58 | 0.61 | 0.58 | 0.54 | 0.02 | 0.04 |
| *Hydromorphone* | 0.18 | 0.19 | 0.15 | 0.14 | 0.12 | 0.12 | 0.13 | 0.10 | 0.08 | -0.10* | -0.57 |
| *Oxycodone* | 1.74 | 1.88 | 2.07 | 2.20 | 2.27 | 2.34 | 2.32 | 2.41 | 2.29 | 0.55* | 0.31 |
| *Dihydrocodeine* | 0.85 | 0.31 | 0.01 | <0.01 | <0.01 | <0.01 | <0.01 | <0.01 | <0.01 | -0.85* | -1.00 |
| *Pethidine* | <0.01 | <0.01 | <0.01 | <0.01 | <0.01 | <0.01 | 0 | 0 | 0 |  |  |
| *Fentanyl* | 2.28 | 2.04 | 2.10 | 2.11 | 2.13 | 2.10 | 2.01 | 1.98 | 1.88 | -0.40* | -0.18 |
| *Buprenorphine* | 1.12 | 1.20 | 1.28 | 1.36 | 1.46 | 1.49 | 1.48 | 1.52 | 1.49 | 0.37* | 0.33 |
| *Tramadol* | 8.83 | 9.20 | 9.45 | 9.38 | 9.29 | 9.09 | 8.80 | 8.91 | 8.48 | -0.35 | -0.04 |
| *Meptazinol* | 0.06 | 0.06 | 0.06 | 0.08 | 0.07 | 0.07 | 0.07 | 0.07 | 0.07 | 0.01 | 0.10 |
| *Tapentadol* | 0.39 | 0.58 | 0.87 | 1.14 | 1.33 | 1.48 | 1.57 | 1.68 | 1.66 | 1.27* | 3.26 |
| *Codeine* | 17.05 | 18.43 | 19.45 | 19.93 | 21.00 | 21.99 | 22.57 | 23.86 | 23.81 | 6.76* | 0.40 |
| Systemic NSAIDs | 47.68 | 48.95 | 49.57 | 49.40 | 49.94 | 49.90 | 48.43 | 50.63 | 49.47 | 1.79 | 0.04 |
| *Non-selective NSAIDs* | 38.28 | 39.18 | 39.63 | 39.44 | 39.95 | 40.08 | 38.66 | 40.60 | 39.83 | 1.54 | 0.04 |
| *Coxib* | 9.40 | 9.78 | 9.94 | 9.96 | 9.99 | 9.82 | 9.77 | 10.03 | 9.64 | 0.25 | 0.03 |
| Paracetamol (incl. combinations) | 51.62 | 55.18 | 58.67 | 62.07 | 66.45 | 69.93 | 73.76 | 78.19 | 79.06 | 27.44* | 0.53 |
| *Paracetamol (excl. opioid combinations)* | 33.26 | 36.33 | 38.74 | 41.40 | 44.61 | 47.04 | 50.20 | 53.30 | 54.00 | 20.75* | 0.62 |
| Gabapentin | 1.89 | 2.01 | 2.19 | 2.36 | 2.53 | 2.70 | 2.89 | 3.14 | 3.29 | 1.40* | 0.74 |
| Pregabalin | 12.91 | 14.58 | 16.04 | 16.78 | 17.11 | 16.92 | 16.47 | 16.93 | 16.73 | 3.82 | 0.30 |
| Low-dose amitriptyline | 0.54 | 0.67 | 0.82 | 1.01 | 1.15 | 1.32 | 1.47 | 1.64 | 1.79 | 1.25* | 2.32 |
| Antimigraines | 1.54 | 1.64 | 1.76 | 1.89 | 1.82 | 1.31 | 1.15 | 1.26 | 1.28 | -0.26 | -0.17 |

\*Test for linear trend significant at $p < 0.05$
<sup>†</sup>Morphine, hydromorphone, oxycodone, pethidine, fentanyl, buprenorphine, tramadol, tapentadol

**Table 2b.** Rate of daily defined doses (DDDs) per 1,000 NHS population per day in England.

|  | 2014 | 2015 | 2016 | 2017 | 2018 | 2019 | 2020 | 2021 | 2022 | Absolute change | Relative change |
| --- | --- | --- | --- | --- | --- | --- | --- | --- | --- | --- | --- |
| All analgesics | 96.96 | 97.81 | 97.23 | 93.93 | 91.00 | 88.36 | 87.65 | 86.58 | 80.56 | -16.40* | -0.17 |
| Opioids | 39.14 | 39.42 | 39.42 | 38.68 | 37.66 | 36.70 | 36.25 | 35.13 | 31.91 | -7.23* | -0.18 |
| *Strong opioids†* | 12.03 | 11.82 | 11.68 | 11.18 | 10.39 | 9.82 | 9.34 | 8.85 | 7.79 | -4.24* | -0.35 |
| *Long-acting opioids* | 5.91 | 5.94 | 5.92 | 5.74 | 5.41 | 5.12 | 4.82 | 4.51 | 3.95 | -1.96* | -0.33 |
| *Morphine* | 2.21 | 2.30 | 2.37 | 2.34 | 2.22 | 2.15 | 2.04 | 1.92 | 1.67 | -0.54* | -0.25 |
| *Hydromorphone* | 0.01 | 0.01 | 0.01 | 0.01 | 0.01 | 0.01 | <0.01 | <0.01 | <0.01 | 0.01* | -0.58 |
| *Oxycodone* | 0.91 | 0.94 | 0.98 | 0.99 | 0.96 | 0.92 | 0.88 | 0.83 | 0.78 | -0.12 | -0.13 |
| *Dihydrocodeine* | 2.97 | 2.80 | 2.63 | 2.45 | 2.29 | 2.13 | 2.02 | 1.90 | 1.66 | -1.31* | -0.44 |
| *Pethidine* | 0.01 | 0.01 | 0.01 | 0.01 | 0.01 | 0.01 | <0.01 | <0.01 | <0.01 | -0.01* | -0.80 |
| *Fentanyl* | 1.23 | 1.25 | 1.24 | 1.18 | 1.08 | 0.98 | 0.89 | 0.79 | 0.66 | -0.57* | -0.47 |
| *Buprenorphine* | 0.67 | 0.70 | 0.73 | 0.75 | 0.75 | 0.75 | 0.74 | 0.73 | 0.67 | -0.01 | -0.01 |
| *Tramadol* | 6.95 | 6.55 | 6.27 | 5.82 | 5.27 | 4.91 | 4.69 | 4.47 | 3.92 | -3.03* | -0.44 |
| *Meptazinol* | 0.05 | 0.05 | 0.05 | 0.04 | 0.04 | 0.04 | 0.04 | 0.04 | 0.03 | -0.01* | -0.26 |
| *Tapentadol* | 0.04 | 0.06 | 0.07 | 0.08 | 0.09 | 0.10 | 0.10 | 0.10 | 0.09 | 0.05* | 1.24 |
| *Codeine* | 24.09 | 24.75 | 25.07 | 25.01 | 24.94 | 24.70 | 24.85 | 24.34 | 22.43 | -1.67 | -0.07 |
| Systemic NSAIDs | 24.76 | 24.09 | 22.88 | 20.77 | 19.36 | 18.57 | 17.54 | 18.01 | 17.76 | -7.00* | -0.28 |
| *Non-selective NSAIDs* | 23.52 | 22.89 | 21.69 | 19.65 | 18.25 | 17.40 | 16.33 | 16.76 | 16.46 | -7.07 | -0.30 |
| *Coxib* | 1.24 | 1.20 | 1.19 | 1.12 | 1.11 | 1.17 | 1.21 | 1.25 | 1.31 | 0.07* | 0.05 |
| Paracetamol (incl. opioid combinations) | 21.43 | 21.25 | 20.61 | 19.06 | 17.76 | 16.67 | 17.02 | 16.21 | 14.57 | -6.86* | -0.32 |
| *Paracetamol (excl. opioid combinations)* | 20.71 | 20.52 | 19.88 | 18.34 | 17.05 | 15.97 | 16.32 | 15.53 | 13.94 | -6.77* | -0.33 |
| Gabapentin | 3.90 | 4.46 | 4.95 | 5.36 | 5.50 | 5.27 | 5.30 | 5.20 | 4.69 | 0.80 | 0.20 |
| Pregabalin | 4.36 | 5.03 | 5.65 | 6.22 | 6.77 | 7.02 | 7.39 | 7.62 | 7.11 | 2.75* | 0.63 |
| Low-dose amitriptyline | 2.28 | 2.44 | 2.57 | 2.67 | 2.75 | 2.86 | 3.01 | 3.15 | 3.23 | 0.95* | 0.42 |
| Antimigraines | 1.80 | 1.85 | 1.87 | 1.89 | 1.91 | 2.56 | 1.84 | 1.94 | 1.91 | 0.11* | 0.06 |

\*Test for linear trend significant at $p < 0.05$
†Morphine, hydromorphone, oxycodone, pethidine, fentanyl, buprenorphine, tramadol, tapentadol

In Ireland, the rate of Oral Morphine Equivalents (OMEs) dispensed per 1,000 GMS population per day for all opioids increased from 1409.8 to 1737.8. The largest increase was seen for tapentadol followed by codeine, while hydromorphone decreased the most. In England, OME rate for opioids decreased from 1262.8 to 911.6. All individual drugs decreased, with the exception of buprenorphine and tapentadol. See Tables 3a and 3b. The rate ratios for OMEs (see Supplementary Table 6 and supplementary Figure 2) were similar to that for dispensings but were of a smaller magnitude, with the rate of OMEs 1.12 times higher in Ireland in 2014, and 1.91 times higher in 2022.

**Table 3a.** Rate of oral morphine equivalent (OME) per 1,000 GMS population per day in Ireland.

|  | 2014 | 2015 | 2016 | 2017 | 2018 | 2019 | 2020 | 2021 | 2022 | Absolute change | Relative change |
| --- | --- | --- | --- | --- | --- | --- | --- | --- | --- | --- | --- |
| Opioids | 1409.81 | 1501.38 | 1607.60 | 1677.12 | 1733.05 | 1768.60 | 1760.74 | 1801.02 | 1737.83 | 328.02* | 0.23 |
| *Strong opioids<sup>†</sup>* | 1224.28 | 1310.09 | 1410.63 | 1475.06 | 1520.53 | 1546.04 | 1532.50 | 1560.00 | 1497.29 | 273.02* | 0.22 |
| *Long-acting opioids* | 690.72 | 734.46 | 797.90 | 845.43 | 877.42 | 897.39 | 887.25 | 896.33 | 863.50 | 172.79* | 0.25 |
| *Morphine* | 50.95 | 52.32 | 54.57 | 54.55 | 54.98 | 57.12 | 59.91 | 56.97 | 52.92 | 1.97 | 0.04 |
| *Hydromorphone* | 13.75 | 15.08 | 11.97 | 10.40 | 9.99 | 10.50 | 11.44 | 8.97 | 7.38 | -6.37* | -0.46 |
| *Oxycodone* | 193.40 | 209.68 | 230.57 | 244.48 | 252.35 | 259.62 | 258.19 | 267.38 | 254.03 | 60.62* | 0.31 |
| *Dihydrocodeine* | 12.81 | 4.72 | 0.21 | 0.05 | 0.01 | 0.01 | 0.01 | <0.001 | <0.001 | -12.81* | -1.00 |
| *Pethidine* | 0.09 | 0.12 | 0.21 | 0.21 | 0.23 | 0.07 | 0 | 0 | 0 |  |  |
| *Fentanyl* | 251.63 | 256.13 | 265.90 | 270.09 | 273.00 | 273.04 | 261.10 | 256.45 | 245.31 | -6.32 | -0.03 |
| *Buprenorphine* | 122.68 | 131.70 | 140.81 | 149.74 | 160.11 | 163.83 | 162.89 | 167.44 | 163.64 | 40.96* | 0.33 |
| *Tramadol* | 529.53 | 552.09 | 567.05 | 562.93 | 557.19 | 545.50 | 528.18 | 534.48 | 508.73 | -20.81 | -0.04 |
| *Meptazinol* | 2.21 | 2.27 | 2.27 | 2.73 | 2.50 | 2.63 | 2.50 | 2.39 | 2.43 | 0.22 | 0.10 |
| *Tapentadol* | 62.24 | 92.96 | 139.56 | 182.66 | 212.68 | 236.36 | 250.80 | 268.31 | 265.29 | 203.05* | 3.26 |
| *Codeine* | 170.51 | 184.30 | 194.50 | 199.29 | 210.00 | 219.92 | 225.73 | 238.62 | 238.10 | 67.59* | 0.40 |

\*Test for trend significant at $p < 0.05$
<sup>†</sup>Morphine, hydromorphone, oxycodone, pethidine, fentanyl, buprenorphine, tramadol, tapentadol

**Table 3b:**
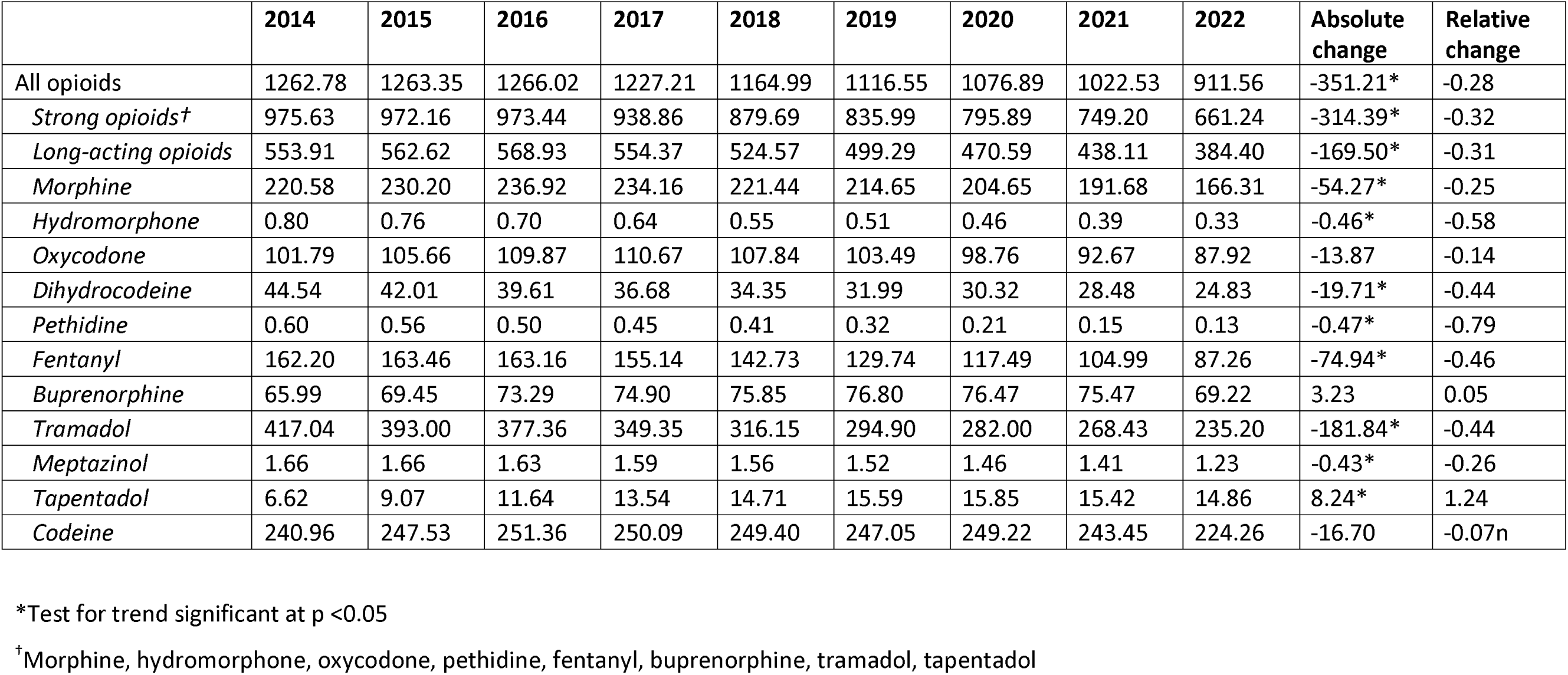
Rate of oral morphine equivalent (OME) per 1,000 NHS population per day in England.

The cost rates decreased in both Ireland and England during the study period. In Ireland the cost rate per 1,000 population decreased by 19%, while the rate in England decreased by 49%. Cost rates for opioids decreased by 39% in England, while rates remained stable in Ireland. Large increases and decreases were seen for individual drugs in both countries. In Ireland, the largest increases were seen for tapentadol (333%), topical NSAIDs (79%), and codeine (60%), while the cost per 1,000 population decreased the most for dihydrocodeine (>99%), lidocaine (84%), and hydromorphone (52%). In England all cost rates decreased, with the exception of tapentadol (126% increase), antimigraines (33%), and low-dose amitriptyline (15%). Adjusted for exchange rate and purchasing power ratio (based on World Bank figures(The World Bank, 2024)), the rate ratio for costs in Ireland versus England was 3.98 in 2014, increasing to 6.52 in 2022. See Supplementary Tables 6 and 7 and Supplementary Figure 2.

Results of the subgroup analysis of the CCGs in England in the top 33 centiles for deprivation showed dispensing rates higher than those of the general population (2083.26 vs 1824.02 in 2014 and 1934.72 vs 1516.54 in 2022 for all analgesics), and thus closer to the rates in Ireland (3388.64 in 2014 and 4316.04 in 2022), although still substantially lower. Similar patterns were seen for most individual drugs. The ratio of dispensing rates between Ireland and the more deprived CCGs are shown in Figure 4. See Supplementary Table 8 for analysis of other outcomes for this deprived subgroup.

**Figure 4.**
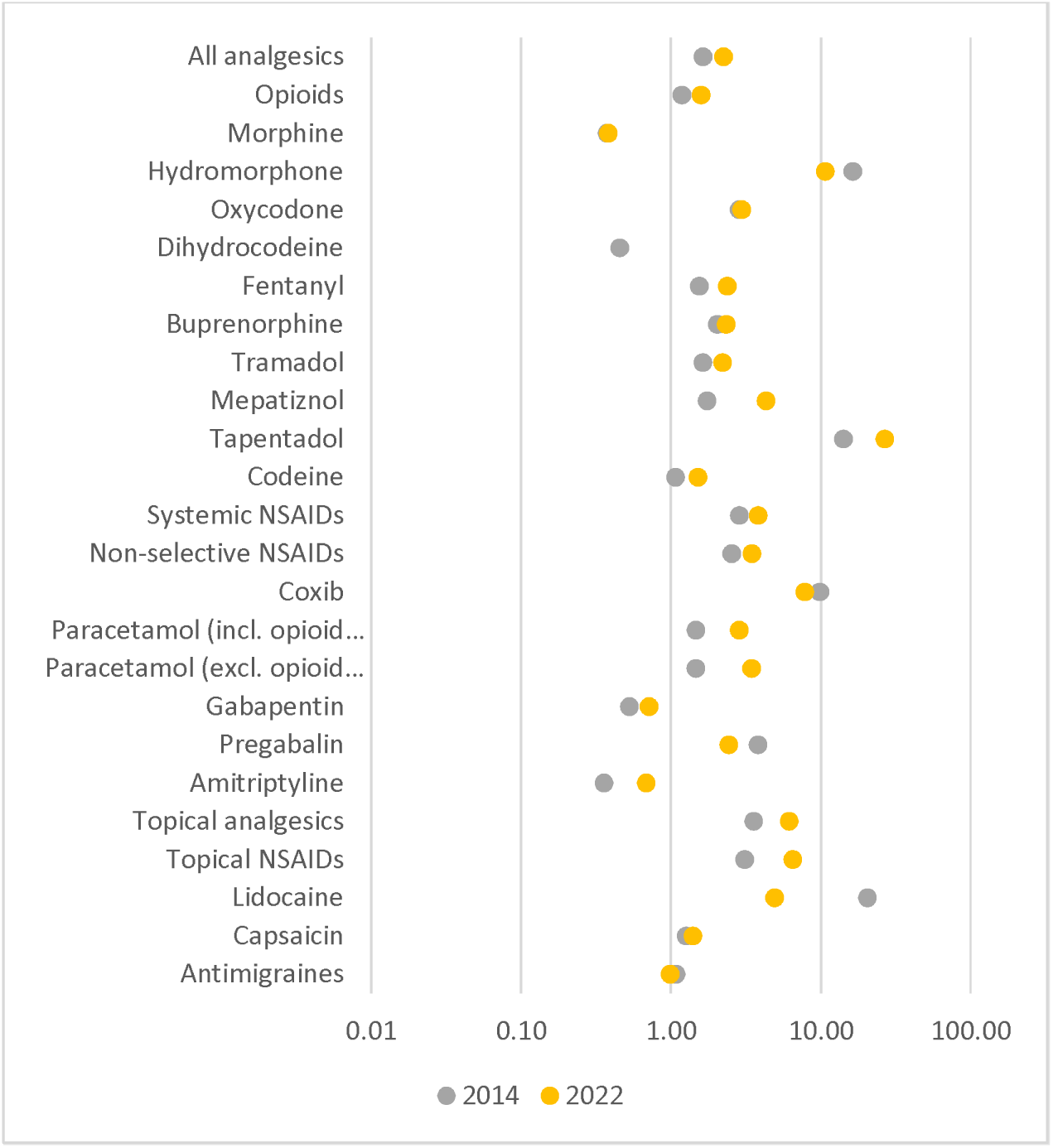
Rate ratio for dispensings in Ireland versus top 33% deprived CCGs in England for 2014 and 2022* *Rates above 1.0 indicate higher dispensings in Ireland Note: Rate ratio for dihydrocodeine dispensings in 2022 was <0.01

## Discussion

In this study, substantially different patterns and trends in analgesia prescribing in Ireland and England between 2014 and 2022 were identified, with higher dispensing rates in Ireland compared to England for most drugs. In the GMS population in Ireland, the rate of overall analgesia and opioids dispensings increased, as well as most individual analgesics. Conversely, the overall analgesia and opioid dispensing rates decreased in England, with similar patterns for most individual drugs. These patterns were consistent when accounting for the strength and quantity of medications dispensing in analysis of DDDs and OMEs, suggesting the changes reflect real shifts in total analgesic consumption. This is in line with previous research focusing on these countries. One recent study found an overall increase in strong opioid prescribing in Ireland between 2010 and 2019, particularly in older adults(Norris et al., 2021). In England, opioid prescriptions have been identified as increasing between 1998 and 2016, before declining in 2017 and 2018(Curtis et al., 2019), and our research shows that this decline has continued since then. Western and Central Europe overall prescribing rates have been found to have increased by 1.6% per year between 2015 and 2019, with rates significantly increasing in some countries (including Ireland) and decreasing in others (including the UK)(Ju et al., 2022).

An important potential reason for the differences in dispensing rates between countries is the difference between the two populations. The NHS rates includes all medications dispensed across all CCGs in England. For Ireland, only medications dispensed to patient eligible for the GMS scheme are included. As outlined above, the GMS scheme is means-tested with older people over-represented. Previous research has identified higher prevalence of pain among older adults and individuals from disadvantaged backgrounds(Yong et al., 2022). A recent study on opioid prescribing in England found a strong association of socioeconomic deprivation with opioid prescribing in primary care(Nowakowska et al., 2021). In that context, it is therefore likely that the GMS population in Ireland would show higher rates of analgesia prescribing compared to the general population in England. The rate ratios of dispensings for Ireland versus England also increased between 2014 and 2022. This may be partly explained by a change in population in the two countries. In Ireland, the GMS population decreased between 2014 and 2022 (as per Supplementary table 3) with GMS eligibility having been shown to have been decreasing in recent years, (Mattsson, Flood, et al., 2022), with the potential implication that the GMS population has become comparatively more deprived, thus increasing analgesia rates. Conversely, the NHS population increased during the study period, primarily driven by immigration(The Migration Observatory, 2023), which is typically a younger population less likely to be prescribed analgesia. To account for differences in deprivation (albeit not in age), we repeated our analysis in the one third of NHS CCGs that are most deprived according to IMD 2019. This analysis found higher dispensing rates compared to the overall NHS population in England, but rates were still substantially lower than the GMS rates in Ireland.

Other reasons for the inter-country differences in utilisation may include universal access in England to health services, including pain clinics and non-pharmacological interventions to manage pain. In Ireland, access to these treatments for GMS patients is often limited. Additionally, waiting times for GMS patients are considerable, in particular for orthopaedics, resulting in patients with severe degenerative related chronic pain potentially waiting several years to be considered for joint replacement surgery, thus requiring strong analgesics in the meantime. Additionally, guidelines on prescribing vary between the health systems in England and Ireland, as well as in some cases the reimbursement systems. In England, guidance on items which should not routinely be prescribed in primary care(NHS England, 2023), specifically addresses oxycodone/naloxone combination products, paracetamol/tramadol combination products, immediate-release fentanyl, and lidocaine plasters. Similar guidelines do not exist in Ireland. In Ireland, however, a new reimbursement system for lidocaine plasters was introduced in 2017; with the result of reducing dispensing rates from 148 per 1,000 population in 2016 to 10 in 2018. This is consistent with previous research on the impact of the reimbursement system(Mattsson et al., 2023; Smith et al., 2021).

Tapentadol had a much higher dispensing rate in Ireland than England, a difference which expanded between 2014 and 2022. Similarly, hydromorphone was more commonly dispensed in Ireland, however the gap narrowed during the study period. In Ireland, the National Clinical Guidelines on pharmacological management of cancer pain in adults outline morphine, hydromorphone, and oxycodone as first-line choices for moderate to severe pain, with no specific guidelines relating to the use of tapentadol, and codeine and codeine/paracetamol combinations as preferable to tramadol or tapentadol for mild to moderate pain(National Clinical Effectiveness Committee, 2015). In England, tapentadol is generally considered a third-line option for moderate to severe pain that should only be prescribed when morphine and oxycodone have proven ineffective or intolerable, while hydromorphone is usually only prescribed by pain teams or palliative care(NHS, 2024). This discrepancy difference between guidelines could explain the higher dispensing rates of the drugs in Ireland.

Drugs that decreased in both Ireland and England include systemic NSAIDs and tramadol. NSAIDs have been identified as a high-risk medicine class by the WHO Global Patient Safety Challenge, Medication Without HarmHarm(WHO, 2019). Previous research on NSAID prescribing in England found that non-selective NSAIDs decreased between 2002 and 2010(Y. Chen et al., 2018), a trend which appears to be continuing. Dispensing of tramadol has varied in the last few decades. A recent study found tramadol dispensing in Ireland to be significantly higher in 2015 compared to 2000(Moriarty et al., 2022), however another study has shown dispensing to be decreasing since 2010(Norris et al., 2021). In England, tramadol dispensing increased between 2010 and 2014(T. C. Chen et al., 2018), however the level of dispensing significantly decreased in 2014 when tramadol was classified as a controlled drug substance. The findings of our study suggest a continued decreasing trend since this classification.

In both Ireland and England, an increase in dispensings of drugs used for neuropathic pain (gabapentin, pregabalin, and low-dose amitriptyline) was noted. In England, gabapentinoid dispensing has been shown to be increasing since the late 1990s (Ashworth et al., 2023). In 2019 they were reclassified as controlled drugs, which appears to have had an effect on dispensing. In our study, gabapentin peaked in 2018 before decreasing and plateauing, albeit at a higher rate compared to 2014. Pregabalin continued to increase until 2021 before similarly plateauing. In Ireland gabapentin and pregabalin are currently not classified as controlled drugs, however guidelines on appropriate prescribing of pregabalin have been issued by the Medicines Management Programme (MMP)(Medicines Management Programme, 2020)

After adjusting for exchange rate and purchase power parity, cost rates remained higher in Ireland compared to England. This discrepancy is potentially driven by a generally higher cost of medicines in Ireland, as well as a higher utilisation of branded drugs compared to England (or in the case of amitriptyline, low-dose formulations only being available as an unlicensed/exempt medicinal product). In recent years measures have been introduced in Ireland to reduce medications costs generally, including generic substitution (HPRA, 2024) and the Preferred Drug Initiative (HSE, 2024; Ronald et al., 2018) although neither of these specifically focus on analgesic medications.

The main strength of our study is the data used. Datasets from both countries cover all prescriptions dispensed to the respective populations within the time period. As these data are primarily used for pharmacies to be reimbursed, the accuracy and completeness of the data are high. Our study does however have some important limitations. Firstly, only aggregate data were analysed, and consequently no comparisons could be made between sub-populations, nor could other patterns of utilisation be analysed. Secondly, although the data sources are comprehensive, they are limited to prescribed medications and non-prescription utilisation is therefore not captured. Lastly, as outlined above the two populations included in the study are not directly comparable, however our sub-group analysis focusing on more deprived CCGs in England explores how these differences may explain the results.

The findings of this study highlight the need for more research using individual, detailed dispensing data. Individual level data would enable analysis of specific prescribing patterns, including switching behaviours, long-term and high-level use, high-risk prescribing of analgesics, as well as clinical indication for analgesic prescribing and off-label prescribing. Higher rates in Ireland suggests a need for enhanced availability of non-pharmacological services and interventions to address pain. Additionally, qualitative work to identify and contextualise analgesia use in primary care and the impact of issues such as orthopaedic waiting times and availability of non-pharmacological interventions is an important area to explore. An important aspect of drug utilisation research is the access to data sources. Restrictions to data sources, including charges and lengthy application processes, may be a challenge to timely analysis and should be addressed by data governance policies, with the availability of open data when appropriate being crucial.

## Data Availability

Code and data for the analysis of NHS data will be available from github. Code for the analysis of GMS data will be available from zenodo. GMS data in aggregated form can be requested from the HSE PCRS at https://www.hse.ie/eng/staff/pcrs/pcrs-publications/.

## Acknowledgements

We would like to acknowledge the wider Controlled Drug Prescribing (CDRx) team.

## Author contributions

FM conceived the study. All authors were involved in the design of the study. MM, BMK, and FM collected and curated the data. MM analysed the data and FM validated the data analysis. All authors were involved the interpretation of the analysis results. MM drafted the manuscript, and MF, BMK, EW, FB, CK, MEW, TF, and FM critically revised the manuscript. FM acquired funding for the study.

## Notes

### Competing Interest Statement

The authors have declared no competing interest.

### Funding Statement

This study is funded by the Health Research Board in Ireland (HRB) through the Secondary Data Analysis Projects scheme (CDRx project, PI FM, grant number SDAP-2019-023). The funder had no role in in study design; in the collection, analysis, and interpretation of data; in the writing of this paper; or in the decision to submit this paper for publication. EW is funded by a HRB Emerging Clinician Scientist Award (grant number: ECSA/2020/002). MEW is funded by a HRB Applying Research into Policy and Practice Award (ARPP/2020/004). OpenPrescribing.net is currently funded by NHS England Primary Care and Medicines Analytics Unit. All other Bennett funding information is available at https://www.bennett.ox.ac.uk/

### Author Declarations

RCSI University of Medicine and Health Sciences Research Ethics Committee (ref: REC202201015) of RCSI Dublin and the Health Service Executive (HSE) Reference Research Ethics Committee B (ref: RRECB1022FM) of the Health Service Exceutive gave ethical approval for this work.

