## Supplementary Material for "Trends in analgesia prescribing in primary care in Ireland and England between 2014 and 2022 - a repeated cross-sectional study"

**Supplementary figure 1. Analgesic and opioid dispensing rates by class 2014-2022**

**Analgesic dispensing rates by analgesic class 2014-2022 in Ireland**

Rate of dispensings per 1,000 GMS population

|  | Opioids | Systemic NSAIDs | Paracetamol (excl. opioid combinations) | Topical analgesics | Antimigraines | Other |
| --- | --- | --- | --- | --- | --- | --- |
| 2014 | 978.652 | 781.02 | 734.339 | 442.412 | 45.6527 | 406.564 |
| 2015 | 1011.9 | 779.945 | 780.614 | 499.408 | 47.9055 | 454.09 |
| 2016 | 1066.79 | 780.409 | 821.142 | 560.664 | 50.6143 | 503.736 |
| 2017 | 1098.57 | 760.081 | 862.318 | 594.93 | 53.0171 | 538.212 |
| 2018 | 1142.95 | 761.69 | 926.885 | 496.418 | 53.3779 | 563.155 |
| 2019 | 1177.24 | 749.532 | 969.851 | 524.073 | 48.3244 | 577.234 |
| 2020 | 1178.98 | 702.891 | 1012.52 | 531.079 | 47.258 | 580.538 |
| 2021 | 1233.47 | 740.686 | 1067.24 | 587.909 | 52.1407 | 613.64 |
| 2022 | 1220.16 | 733.667 | 1092.78 | 589.381 | 53.334 | 628.996 |

**Analgesic dispensing rates by analgesic class for 2014-2022 in England**

Rate of dispensings per 1,000 NHS population

|  | Opioids | Systemic NSAIDs | Paracetamol (excl. opioid combinations) | Topical analgesics | Antimigraines | Other |
| --- | --- | --- | --- | --- | --- | --- |
| 2014 | 720.514 | 258.761 | 407.804 | 107.919 | 47.1886 | 281.832 |
| 2015 | 720.687 | 248.904 | 397.707 | 114.011 | 48.5202 | 313.609 |
| 2016 | 720.601 | 233.353 | 380.929 | 120.158 | 49.2864 | 343.996 |
| 2017 | 702.163 | 206.932 | 343.286 | 118.36 | 49.8543 | 368.297 |
| 2018 | 679.703 | 190.266 | 312.208 | 112.4 | 50.6315 | 386.562 |
| 2019 | 665.444 | 180.654 | 286.548 | 99.3218 | 52.5895 | 394.127 |
| 2020 | 628.371 | 166.63 | 286.689 | 70.9672 | 51.6771 | 408.164 |
| 2021 | 642.724 | 170.646 | 268.434 | 71.587 | 54.1703 | 418.726 |
| 2022 | 584.594 | 167.08 | 238.919 | 70.6448 | 53.159 | 402.143 |

**Opioid dispensing rates by drug for 2014-2022 in Ireland**

Rate of dispesings per 1,000 GMS population

| Year | Morphine | Oxycodone | Dihydrocodeine | Fentanyl | Buprenorphine | Tramadol | Tapentadol | Codeine | Other |
| --- | --- | --- | --- | --- | --- | --- | --- | --- | --- |
| 2014 | 29.0066 | 76.2334 | 40.4676 | 34.3727 | 68.7352 | 271.289 | 13.9916 | 439.046 | 5.51083 |
| 2015 | 29.7443 | 83.1125 | 10.5692 | 34.6329 | 75.4462 | 278.76 | 21.7211 | 472.494 | 5.42294 |
| 2016 | 31.3156 | 92.2204 | 0.545198 | 36.185 | 81.5813 | 284.359 | 34.2079 | 501.273 | 5.10039 |
| 2017 | 32.854 | 100.532 | 0.103738 | 36.6501 | 85.8922 | 279.995 | 44.5895 | 512.634 | 5.31985 |
| 2018 | 34.7011 | 105.55 | 0.045366 | 37.5241 | 91.4776 | 275.987 | 52.7242 | 539.952 | 4.99026 |
| 2019 | 37.1976 | 111.225 | 0.0297855 | 37.3284 | 92.5456 | 270.066 | 59.3101 | 564.435 | 5.10692 |
| 2020 | 39.7169 | 109.963 | 0.014513 | 35.919 | 91.3812 | 260.889 | 62.8897 | 573.155 | 5.05114 |
| 2021 | 38.3686 | 118.968 | 0.00517725 | 35.4881 | 93.9522 | 266.122 | 68.6173 | 607.638 | 4.30618 |
| 2022 | 37.2722 | 114.432 | 0.00510081 | 33.9414 | 91.7527 | 256.659 | 68.4509 | 613.83 | 3.81987 |

**Opioid dispensing rates by drug for 2014-2022 in England**

Rate of dispensing per 1,000 NHS population

| Year | Morphine | Oxycodone | Dihydrocodeine | Fentanyl | Buprenorphine | Tramadol | Tapentadol | Codeine | Other |
| --- | --- | --- | --- | --- | --- | --- | --- | --- | --- |
| 2014 | 71.3005 | 23.9001 | 80.7253 | 21.2648 | 33.6197 | 139.54 | 1.14258 | 347.135 | 1.93017 |
| 2015 | 77.0653 | 25.8197 | 75.1475 | 21.4504 | 35.816 | 131.633 | 1.52052 | 350.368 | 1.9022 |
| 2016 | 82.3612 | 27.7496 | 70.1842 | 21.5233 | 38.0149 | 126.631 | 1.97573 | 350.339 | 1.8545 |
| 2017 | 84.4272 | 29.1897 | 64.4477 | 20.437 | 38.7496 | 117.542 | 2.31368 | 343.312 | 1.76999 |
| 2018 | 83.1098 | 29.9239 | 59.6301 | 18.8505 | 38.9736 | 107.114 | 2.51104 | 337.921 | 1.68922 |
| 2019 | 84.6474 | 30.544 | 55.2986 | 17.3534 | 39.7297 | 101.324 | 2.68994 | 332.285 | 1.59367 |
| 2020 | 85.3406 | 30.67 | 51.9002 | 15.9358 | 39.8952 | 98.0616 | 2.74149 | 332.461 | 1.43759 |
| 2021 | 83.7283 | 30.4272 | 48.9475 | 14.4248 | 39.9577 | 94.96 | 2.69088 | 326.277 | 1.32875 |
| 2022 | 75.3329 | 30.1177 | 42.4938 | 12.0751 | 36.9624 | 83.898 | 2.62965 | 299.945 | 1.15579 |

**Supplementary figure 2. Rate ratios for costs, DDDs, and OMEs in Ireland versus England for 2014 and 2022**

**Rate ratios for costs (adjusted for exchange rate and purchase power parity*)**

|  | 2014 | 2022 |
| --- | --- | --- |
| Antimigraines | 2.28238 | 1.96426 |
| Capsaicin | 2.22159 | 3.11722 |
| Lidocaine | 39.9479 | 10.8326 |
| Topical NSAIDs | 7.02986 | 29.0537 |
| Topical analgesics | 15.7382 | 18.9761 |
| Amitriptyline | 1.33497 | 3.85242 |
| Pregabalin | 4.09992 | 16.0621 |
| Gabapentin | 2.54919 | 3.84227 |
| Paracetamol (excl. opioid combinations) | 2.11256 | 5.93451 |
| Paracetamol (incl. opioid combinations) | 2.92249 | 6.48856 |
| Coxib | 10.6178 | 16.1819 |
| Non-selective NSAIDs | 4.79513 | 9.80485 |
| Systemic NSAIDs | 6.16197 | 10.7697 |
| Codeine | 2.35071 | 5.84905 |
| Tapentadol | 13.151 | 25.7227 |
| Meptazinol | 2.19205 | 2.89578 |
| Tramadol | 3.9737 | 8.12865 |
| Buprenorphine | 3.7141 | 4.05366 |
| Fentanyl | 3.503 | 5.44937 |
| Dihydrocodeine | 0.586423 | 0.01 |
| Oxycodone | 2.12787 | 4.5343 |
| Hydromorphone | 21.3647 | 24.3324 |
| Morphine | 0.710193 | 0.900099 |
| Opioids | 2.76364 | 5.08742 |
| All analgesics | 3.97579 | 6.51623 |

|  | 2014 |
| --- | --- |
| 2.28238 | 0.5 |
| 2.22159 | 1.5 |
| 39.9479 | 2.5 |
| 7.02986 | 3.5 |
| 15.7382 | 4.5 |
| 1.33497 | 5.5 |
| 4.09992 | 6.5 |
| 2.54919 | 7.5 |
| 2.11256 | 8.5 |
| 2.92249 | 9.5 |
| 10.6178 | 10.5 |
| 4.79513 | 11.5 |
| 6.16197 | 12.5 |
| 2.35071 | 13.5 |
| 13.151 | 14.5 |
| 2.19205 | 15.5 |
| 3.9737 | 16.5 |
| 3.7141 | 17.5 |
| 3.503 | 18.5 |
| 0.586423 | 19.5 |
| 2.12787 | 20.5 |
| 21.3647 | 21.5 |
| 0.710193 | 22.5 |
| 2.76364 | 23.5 |
| 3.97579 | 24.5 |

|  | 2022 |
| --- | --- |
| 1.96426 | 0.5 |
| 3.11722 | 1.5 |
| 10.8326 | 2.5 |
| 29.0537 | 3.5 |
| 18.9761 | 4.5 |
| 3.85242 | 5.5 |
| 16.0621 | 6.5 |
| 3.84227 | 7.5 |
| 5.93451 | 8.5 |
| 6.48856 | 9.5 |
| 16.1819 | 10.5 |
| 9.80485 | 11.5 |
| 10.7697 | 12.5 |
| 5.84905 | 13.5 |
| 25.7227 | 14.5 |
| 2.89578 | 15.5 |
| 8.12865 | 16.5 |
| 4.05366 | 17.5 |
| 5.44937 | 18.5 |
| 0.01 | 19.5 |
| 4.5343 | 20.5 |
| 24.3324 | 21.5 |
| 0.900099 | 22.5 |
| 5.08742 | 23.5 |
| 6.51623 | 24.5 |

*The World Bank. Word Development Indicators. <https://databank.worldbank.org/reports.aspx?source=2&series=PA.NUS.FCRF&country>=

**Rate ratios for DDDs**

|  | 2014 | 2022 |
| --- | --- | --- |
| Antimigraines | 0.852522 | 0.670998 |
| Amitriptyline | 0.2361 | 0.553841 |
| Pregabalin | 2.96 | 2.35 |
| Gabapentin | 0.48 | 0.7 |
| Paracetamol (excl. opioid combinations) | 1.60617 | 3.87386 |
| Paracetamol (incl. opioid combinations) | 2.40898 | 5.42625 |
| Coxib | 7.56155 | 7.37032 |
| Non-selective NSAIDs | 1.62764 | 2.42036 |
| Systemic NSAIDs | 1.92539 | 2.78497 |
| Codeine | 0.707679 | 1.06174 |
| Tapentadol | 9.39647 | 17.8469 |
| Mepatiznol | 1.33108 | 1.97217 |
| Tramadol | 1.26973 | 2.16294 |
| Buprenorphine | 1.65763 | 2.23506 |
| Fentanyl | 1.85183 | 2.85934 |
| Dihydrocodeine | 0.287643 | 0.01 |
| Oxycodone | 1.9204 | 2.91445 |
| Hydromorphone | 22.8733 | 23.3598 |
| Morphine | 0.233466 | 0.322116 |
| Opioids | 0.84357 | 1.26254 |
| All analgesics | 1.19676 | 1.81987 |

|  | 2014 |
| --- | --- |
| 0.852522 | 0.5 |
| 0.2361 | 1.5 |
| 2.96 | 2.5 |
| 0.48 | 3.5 |
| 1.60617 | 4.5 |
| 2.40898 | 5.5 |
| 7.56155 | 6.5 |
| 1.62764 | 7.5 |
| 1.92539 | 8.5 |
| 0.707679 | 9.5 |
| 9.39647 | 10.5 |
| 1.33108 | 11.5 |
| 1.26973 | 12.5 |
| 1.65763 | 13.5 |
| 1.85183 | 14.5 |
| 0.287643 | 15.5 |
| 1.9204 | 16.5 |
| 22.8733 | 17.5 |
| 0.233466 | 18.5 |
| 0.84357 | 19.5 |
| 1.19676 | 20.5 |

|  | 2022 |
| --- | --- |
| 0.670998 | 0.5 |
| 0.553841 | 1.5 |
| 2.35 | 2.5 |
| 0.7 | 3.5 |
| 3.87386 | 4.5 |
| 5.42625 | 5.5 |
| 7.37032 | 6.5 |
| 2.42036 | 7.5 |
| 2.78497 | 8.5 |
| 1.06174 | 9.5 |
| 17.8469 | 10.5 |
| 1.97217 | 11.5 |
| 2.16294 | 12.5 |
| 2.23506 | 13.5 |
| 2.85934 | 14.5 |
| 0.01 | 15.5 |
| 2.91445 | 16.5 |
| 23.3598 | 17.5 |
| 0.322116 | 18.5 |
| 1.26254 | 19.5 |
| 1.81987 | 20.5 |

**Rate ratios for OMEs**

|  | 2014 | 2022 |
| --- | --- | --- |
| Codeine | 0.707641 | 1.06172 |
| Tapentadol | 9.39647 | 17.8469 |
| Mepatiznol | 1.33108 | 1.97217 |
| Tramadol | 1.26973 | 2.16294 |
| Buprenorphine | 1.85897 | 2.36385 |
| Fentanyl | 1.55132 | 2.81124 |
| Dihydrocodeine | 0.287643 | 0.01 |
| Oxycodone | 1.90005 | 2.88937 |
| Hydromorphone | 17.2241 | 22.114 |
| Morphine | 0.230985 | 0.318227 |
| Longacting opioids | 1.24699 | 2.24635 |
| Strong opioids | 1.25485 | 2.26436 |
| Opioids | 1.11644 | 1.90643 |

|  | 2014 |
| --- | --- |
| 0.707641 | 0.5 |
| 9.39647 | 1.5 |
| 1.33108 | 2.5 |
| 1.26973 | 3.5 |
| 1.85897 | 4.5 |
| 1.55132 | 5.5 |
| 0.287643 | 6.5 |
| 1.90005 | 7.5 |
| 17.2241 | 8.5 |
| 0.230985 | 9.5 |
| 1.24699 | 10.5 |
| 1.25485 | 11.5 |
| 1.11644 | 12.5 |

|  | 2022 |
| --- | --- |
| 1.06172 | 0.5 |
| 17.8469 | 1.5 |
| 1.97217 | 2.5 |
| 2.16294 | 3.5 |
| 2.36385 | 4.5 |
| 2.81124 | 5.5 |
| 0.01 | 6.5 |
| 2.88937 | 7.5 |
| 22.114 | 8.5 |
| 0.318227 | 9.5 |
| 2.24635 | 10.5 |
| 2.26436 | 11.5 |
| 1.90643 | 12.5 |

**Supplementary table 1. Drugs included in the analysis**

| **Medication** | **ATC-code** | **BNF-code** |
| --- | --- | --- |
| **Opioids** |  |  |
| ***Strong opioids (as defined by BNF)^1^*** |  |  |
| Morphine, incl. combinations^2^ | N02AA01,  N02AA51 | 0407020P0,  0407020Q0,  040702020 |
| Hydromorphone^2^ | N02AA03 | 040702050 |
| Oxycodone, incl. combinations^2^ | N02AA05,  N02AA55 | 0407020AD,  0407020AF |
| Pethidine | N02AB02 | 0407020V0 |
| Fentanyl^3^ | N02AB03 | 0407020A0 |
| Buprenorphine^3^ | N02AE01 | 0407020B0 |
| Tramadol^2^ | N02AX02,  N02AJ13,  N02AJ14 | 040702040 |
| Tapentadol^2^ | N02AX06 | 0407020AG |
| ***Weak opioids*** |  |  |
| Meptazinol | N02AX05 | 0407020L0 |
| Dihydrocodeine, incl. combinations | N02AA08,  N02AJ01 | 0407020G0,  0407010N0 |
| Codeine, incl. combinations | R05DA04,  N02AJ06 | 0407020C0,  0407010F0 |
| **Systemic NSAIDs** |  |  |
| ***Coxibs*** |  |  |
| Celecoxib | M01AH01 | 1001010AH |
| Etoricoxib | M01AH05 | 1001010AJ |
| ***Non-selective NSAIDs*** |  |  |
| Indometacin | M01AB01 | 1001010K0 |
| Diclofenac, incl. combinations | M01AB05,  M01AB55 | 1001010AG,  1001010C0 |
| Aceclofenac | M01AB16 | 100101080 |
| Meloxicam | M01AC06 | 1001010AA |
| Ibuprofen | M01AE01 | 1001010J0 |
| Naproxen, incl. combinations | M01AE02,  M01AE52 | 1001010P0 |
| Ketoprofen | M01AE03 | 1001010L0 |
| Flurbiprofen | M01AE09 | 1001010I0 |
| Dexketoprofen | M01AE17,  N02AJ14 | 1001010AE, 040702040 |
| Mefenamic acid | M01AG01 | 1001010N0 |
| Nabumetone | M01AX01 | 1001010X0 |
| **Paracetamol, incl. combinations** | N02BE01,  N02BE51 | 0407010H0,  0407010U0,  0407010X0 |
| **Topical analgesics** |  |  |
| ***Topical NSAIDs*** |  |  |
| Benzydamine | M02AA05 | 1003020F0 |
| Etofenamate | M02AA06 |  |
| Piroxicam | M02AA07 | 1003020R0 |
| Ketoprofen | M02AA10 | 100302010 |
| Ibuprofen | M02AA13 | 1003020P0 |
| Diclofenac | M02AA15 | 1003020AF,  1003020U0 |
| ***Other topical agents*** |  |  |
| Capsaicin | M02AB01 | 1003020AA |
| Lidocaine | N01BB02 | 1502010J0 |
| **Other analgesics** |  |  |
| Gabapentin | N03AX12 | 0408010G0 |
| Pregabalin | N03AX16 | 0408010AE |
| Amitriptyline 10mg | N06AA09 | 0403010B0,  0403010B0AAAGAG |
| **Antimigraine preparations** |  |  |
| Sumatriptan | N02CC01 | 0407041T0 |
| Naratriptan | N02CC02 | 0407041M0 |
| Zolmitriptan | N02CC03 | 0407041Z0 |
| Rizatriptan | N02CC04 | 0407041R0 |
| Almotriptan | N02CC05 | 0407041B0 |
| Eletriptan | N02CC06 | 0407041AA |
| Frovatriptan | N02CC07 | 0407041AB |
| Clonidine | N02CX02 | 0407042F0 |

^1^Joint Formulary Committee. British national formulary 78. British Medical Association and Royal Pharmaceutical Society of Great Britain; 2024

^2^Including long-acting oral formulations

^3^Including long-acting transdermal patches

**Supplementary table 2. Oral morphine equivalents (OME)**

| **Drug** | **OME equivalent** | **Source** |
| --- | --- | --- |
| Morphine (oral) | 1 | Nielsen et al^1^ |
| Morphine (rectal) | 1 | Not available, assumed same as oral |
| Morphine (parenteral) | 3 | Nielsen et al |
| Hydromorphone (oral) | 5 | Nielsen et al |
| Hydromorphone (parenteral) | 17.5 | Nielsen et al |
| Oxycodone (oral) | 1.5 | Nielsen et al |
| Oxycodone (parenteral) | 3 | Nielsen et al |
| Dihydrocodeine | 0.1 | Nielsen et al |
| Pethidine (oral) | 0.1 | Nielsen et al |
| Pethidine (parenteral) | 0.4 | Nielsen et al |
| Fentanyl (oral) | 100 | Nielsen et al |
| Fentanyl (nasal) | 160 | Curtis et al^2^ |
| Fentanyl (transdermal) | 112.5 | Nielsen et al |
| Buprenorphine (oral) | 38.8 | Nielsen et al |
| Buprenorphine (parenteral) | 75 | Nielsen et al |
| Buprenorphine (transdermal) | 91.67 | Nielsen et al |
| Tramadol | 0.2 | Nielsen et al |
| Meptazinol | 0.03 | Curtis et al |
| Tapentadol | 0.4 | Nielsen et al |
| Codeine (oral) | 0.1 | Nielsen et al |
| Codeine (parenteral) | 0.3 | Nielsen et al |

1.	Nielsen S, Degenhardt L, Hoban B, Gisev N. A synthesis of oral morphine equivalents (OME) for opioid utilisation studies. *Pharmacoepidemiology and Drug Safety*. 2016;25(6):733-737. doi:<https://doi.org/10.1002/pds.3945>

2.	Curtis HJ, Croker R, Walker AJ, Richards GC, Quinlan J, Goldacre B. Opioid prescribing trends and geographical variation in England, 1998&#x2013;2018: a retrospective database study. *The Lancet Psychiatry*. 2019;6(2):140-150. doi:10.1016/S2215-0366(18)30471-1

**Supplementary table 3. GMS (Ireland) and NHS (England) populations**

|  | **GMS** | **NHS** |
| --- | --- | --- |
| **2014** | 1,768,700 | 56,545,892 |
| **2015** | 1,734,853 | 57,111,235 |
| **2016** | 1,683,792 | 57,744,814 |
| **2017** | 1,609,820 | 58,492,541 |
| **2018** | 1,565,049 | 59,178,163 |
| **2019** | 1,544,374 | 59,901,236 |
| **2020** | 1,584,790 | 60,413,787 |
| **2021** | 1,545,222 | 60,970,002 |
| **2022** | 1,568,379 | 61,768,942 |

**Supplementary Table 4. Absolute dispensings for Ireland and England**

**Absolute dispensings for GMS (Ireland)**

|  | **2014** | **2015** | **2016** | **2017** | **2018** | **2019** | **2020** | **2021** | **2022** | **Absolute change** | **Relative change** |
| --- | --- | --- | --- | --- | --- | --- | --- | --- | --- | --- | --- |
| **All analgesics** | 5993487 | 6200132 | 6370379 | 6289772 | 6173177 | 6246122 | 6420336 | 6633690 | 6769182 | 775695 | 0.13 |
| **Opioids** | 1730942 | 1755503 | 1796249 | 1768500 | 1788776 | 1818106 | 1868435 | 1905978 | 1913678 | 182736 | 0.11 |
| ***Strong opioids*** | 879282 | 913956 | 947814 | 938878 | 939807 | 942236 | 956066 | 963430 | 947373 | 68091 | 0.08 |
| ***Longacting opioids*** | 359381 | 377523 | 399815 | 404011 | 412164 | 415286 | 420416 | 420319 | 414530 | 55149 | 0.15 |
| ***Morphine*** | 51304 | 51602 | 52729 | 52889 | 54309 | 57447 | 62943 | 59288 | 58457 | 7153 | 0.14 |
| ***Hydromorphone*** | 5957 | 5597 | 4790 | 4148 | 3776 | 3702 | 3989 | 3049 | 2412 | -3545 | -0.60 |
| ***Oxycodone*** | 134834 | 144188 | 155280 | 161839 | 165191 | 171773 | 174268 | 183832 | 179473 | 44639 | 0.33 |
| ***Dihydrocodeine*** | 71575 | 18336 | 918 | 167 | 71 | 46 | 23 | 8 | 8 | -71567 | -1.00 |
| ***Pethidine*** | 245 | 308 | 320 | 209 | 188 | 60 | 0 | 0 | 0 | -245 | -1.00 |
| ***Fentanyl*** | 60795 | 60083 | 60928 | 59000 | 58727 | 57649 | 56924 | 54837 | 53233 | -7562 | -0.12 |
| ***Buprenorphine*** | 121572 | 130888 | 137366 | 138271 | 143167 | 142925 | 144820 | 145177 | 143903 | 22331 | 0.18 |
| ***Tramadol*** | 479828 | 483607 | 478802 | 450741 | 431933 | 417083 | 413455 | 411218 | 402538 | -77290 | -0.16 |
| ***Meptazinol*** | 3545 | 3503 | 3478 | 4207 | 3846 | 4125 | 4016 | 3605 | 3579 | 34 | 0.01 |
| ***Tapentadol*** | 24747 | 37683 | 57599 | 71781 | 82516 | 91597 | 99667 | 106029 | 107357 | 82610 | 3.34 |
| ***Codeine*** | 776540 | 819708 | 844039 | 825248 | 845052 | 871699 | 908330 | 938935 | 962718 | 186178 | 0.24 |
| **Systemic NSAIDs** | 1381390 | 1353090 | 1314046 | 1223593 | 1192082 | 1157558 | 1113934 | 1144525 | 1150668 | -230722 | -0.17 |
| ***Non-selective NSAIDs*** | 1177308 | 1146955 | 1110933 | 1026875 | 1001141 | 973068 | 926202 | 955624 | 964659 | -212649 | -0.18 |
| ***Coxib*** | 204082 | 206135 | 203113 | 196718 | 190941 | 184490 | 187732 | 188901 | 186009 | -18073 | -0.09 |
| **Paracetamol (incl. combinations)** | 2289705 | 2345494 | 2393188 | 2377839 | 2457206 | 2530178 | 2680494 | 2764928 | 2860431 | 570726 | 0.25 |
| ***Paracetamol (excl. combinations)*** | 1298825 | 1354251 | 1382632 | 1388176 | 1450620 | 1497812 | 1604637 | 1649119 | 1713899 | 415074 | 0.32 |
| **Gabapentin** | 96530 | 100972 | 108169 | 115040 | 122876 | 131207 | 144531 | 157157 | 168927 | 72397 | 0.75 |
| **Pregabalin** | 547081 | 597374 | 636544 | 633526 | 626628 | 612814 | 610617 | 614739 | 620485 | 73404 | 0.13 |
| **Amitriptyline** | 75479 | 89433 | 103474 | 117859 | 131861 | 147444 | 164883 | 176314 | 197092 | 121613 | 1.61 |
| **Topical analgesics** | 782494 | 866400 | 944041 | 957730 | 776919 | 809364 | 841648 | 908450 | 924373 | 141879 | 0.18 |
| ***Topical NSAIDs*** | 631999 | 661189 | 683185 | 708877 | 743070 | 774076 | 811622 | 875116 | 891545 | 259546 | 0.41 |
| ***Lidocaine*** | 138957 | 193364 | 248980 | 235349 | 16330 | 21886 | 23081 | 21477 | 20418 | -118539 | -0.85 |
| ***Capsaicin*** | 11538 | 11847 | 11876 | 13504 | 17519 | 13402 | 6945 | 11857 | 12410 | 872 | 0.08 |
| **Antimigraines** | 80746 | 83109 | 85224 | 85348 | 83539 | 74631 | 74894 | 80569 | 83648 | 2902 | 0.04 |

**Absolute dispensings for NHS (England)**

|  | **2014** | **2015** | **2016** | **2017** | **2018** | **2019** | **2020** | **2021** | **2022** | **Absolute change** | **Relative change** |
| --- | --- | --- | --- | --- | --- | --- | --- | --- | --- | --- | --- |
| **All analgesics** | 103140742 | 105280888 | 106731075 | 104636801 | 102482961 | 100555207 | 99232713 | 99154818 | 93675104 | -9465639 | -0.09 |
| **Opioids** | 40742109 | 41159311 | 41610954 | 41071282 | 40223575 | 39860913 | 37962296 | 39186902 | 36109760 | -4632349 | -0.11 |
| ***Strong opioids*** | 16484612 | 16791702 | 17260554 | 17153152 | 16630084 | 16576415 | 14674796 | 16244526 | 14900776 | -1583835 | -0.10 |
| ***Longacting opioids*** | 7452792 | 7831800 | 8177786 | 8266590 | 8148866 | 8207242 | 7066782 | 7950345 | 7295889 | -156903 | -0.02 |
| ***Morphine*** | 4031749 | 4401295 | 4755935 | 4938360 | 4918286 | 5070481 | 5155748 | 5104917 | 4653233 | 621484 | 0.15 |
| ***Hydromorphone*** | 9680 | 9524 | 9407 | 9761 | 8676 | 8425 | 7619 | 6857 | 6336 | -3344 | -0.35 |
| ***Oxycodone*** | 1351455 | 1474597 | 1602398 | 1707381 | 1770840 | 1829623 | 1852891 | 1855149 | 1860338 | 508883 | 0.38 |
| ***Dihydrocodeine*** | 4564683 | 4291768 | 4052774 | 3769707 | 3528798 | 3312455 | 3135489 | 2984328 | 2624799 | -1939884 | -0.42 |
| ***Pethidine*** | 33193 | 31165 | 28410 | 25004 | 22963 | 17943 | 11273 | 8123 | 7157 | -26036 | -0.78 |
| ***Fentanyl*** | 1202437 | 1225061 | 1242858 | 1195411 | 1115536 | 1039493 | 962743 | 879479 | 745865 | -456572 | -0.38 |
| ***Buprenorphine*** | 1901057 | 2045497 | 2195163 | 2266560 | 2306389 | 2379858 | 2410219 | 2436224 | 2283126 | 382069 | 0.20 |
| ***Tramadol*** | 7890433 | 7517724 | 7312295 | 6875343 | 6338796 | 6069461 | 5924274 | 5789713 | 5182291 | -2708142 | -0.34 |
| ***Meptazinol*** | 66270 | 67948 | 69271 | 68766 | 68326 | 69095 | 67958 | 66034 | 57899 | -8371 | -0.13 |
| ***Tapentadol*** | 64608 | 86839 | 114088 | 135333 | 148599 | 161131 | 165624 | 164063 | 162431 | 97823 | 1.51 |
| ***Codeine*** | 19629052 | 20009964 | 20230276 | 20081208 | 19997563 | 19904292 | 20085200 | 19893117 | 18527301 | -1101751 | -0.06 |
| **Systemic NSAIDs** | 14631885 | 14215219 | 13474911 | 12103968 | 11259604 | 10821409 | 10066771 | 10404317 | 10320345 | -4311540 | -0.29 |
| ***Non-selective NSAIDs*** | 13971340 | 13572733 | 12829299 | 11472840 | 10619746 | 10129325 | 9338557 | 9631391 | 9501656 | -4469684 | -0.32 |
| ***Coxib*** | 660545 | 642486 | 645612 | 631128 | 639858 | 692084 | 728214 | 772926 | 818688 | 158143 | 0.24 |
| **Paracetamol (incl. combinations)** | 41479584 | 40995570 | 40038324 | 37598551 | 35605981 | 33920830 | 34079652 | 32774491 | 29917260 | -11562324 | -0.28 |
| ***Paracetamol (excl. combinations)*** | 23059628 | 22713526 | 21996700 | 20079666 | 18475891 | 17164550 | 17319957 | 16366443 | 14757792 | -8301836 | -0.36 |
| **Gabapentin** | 4919912 | 5696090 | 6435279 | 7063120 | 7346854 | 7243460 | 7386138 | 7343571 | 6762478 | 1842566 | 0.37 |
| **Pregabalin** | 4031674 | 4752330 | 5494505 | 6206353 | 6957889 | 7367932 | 7821306 | 8246432 | 7824047 | 3792373 | 0.94 |
| **Amitriptyline** | 6984840 | 7462154 | 7934202 | 8273128 | 8571261 | 8997274 | 9451295 | 9939726 | 10253448 | 3268608 | 0.47 |
| **Topical analgesics** | 6102372 | 6511304 | 6938488 | 6923178 | 6651600 | 5949499 | 4287395 | 4364659 | 4363657 | -1738715 | -0.28 |
| ***Topical NSAIDs*** | 5667149 | 6037029 | 6395403 | 6372769 | 6144591 | 5584608 | 3952880 | 3954595 | 3995759 | -1671390 | -0.29 |
| ***Lidocaine*** | 190067 | 223840 | 254566 | 250935 | 198963 | 158834 | 129990 | 122739 | 121138 | -68929 | -0.36 |
| ***Capsaicin*** | 245156 | 250435 | 288519 | 299474 | 308046 | 206057 | 204525 | 287325 | 246760 | 1604 | 0.01 |
| **Antimigraines** | 2668323 | 2771051 | 2846036 | 2916105 | 2996280 | 3150173 | 3122011 | 3302764 | 3283573 | 615250 | 0.23 |

**Supplementary table 5. Proportional share of analgesics and opioids for Ireland and England**

| **Proportional share of analgesic prescribing (Ireland)** | | | | | | | | | |
| --- | --- | --- | --- | --- | --- | --- | --- | --- | --- |
|  | **2014** | **2015** | **2016** | **2017** | **2018** | **2019** | **2020** | **2021** | **2022** |
| Opioids | 0.29 | 0.28 | 0.28 | 0.28 | 0.29 | 0.29 | 0.29 | 0.29 | 0.28 |
| *Strong opioids* | 0.15 | 0.15 | 0.15 | 0.15 | 0.15 | 0.15 | 0.15 | 0.15 | 0.14 |
| *Long-acting opioids* | 0.06 | 0.06 | 0.06 | 0.06 | 0.07 | 0.07 | 0.07 | 0.06 | 0.06 |
| *Morphine* | 0.01 | 0.01 | 0.01 | 0.01 | 0.01 | 0.01 | 0.01 | 0.01 | 0.01 |
| *Hydromorphone* | <0.01 | <0.01 | <0.01 | <0.01 | <0.01 | <0.01 | <0.01 | <0.01 | <0.01 |
| *Oxycodone* | 0.02 | 0.02 | 0.02 | 0.03 | 0.03 | 0.03 | 0.03 | 0.03 | 0.03 |
| *Dihydrocodeine* | 0.01 | <0.01 | <0.01 | <0.01 | <0.01 | <0.01 | <0.01 | <0.01 | <0.01 |
| *Pethidine* | <0.01 | <0.01 | <0.01 | <0.01 | <0.01 | <0.01 | 0 | 0 | 0 |
| *Fentanyl* | 0.01 | 0.01 | 0.01 | 0.01 | 0.01 | 0.01 | 0.01 | 0.01 | 0.01 |
| *Buprenorphine* | 0.02 | 0.02 | 0.02 | 0.02 | 0.03 | 0.03 | 0.03 | 0.02 | 0.02 |
| *Tramadol* | 0.08 | 0.08 | 0.08 | 0.07 | 0.07 | 0.07 | 0.06 | 0.06 | 0.06 |
| *Meptazinol* | <0.01 | <0.01 | <0.01 | <0.01 | <0.01 | <0.01 | <0.01 | <0.01 | <0.01 |
| *Tapentadol* | <0.01 | 0.01 | 0.01 | 0.01 | 0.01 | 0.01 | 0.02 | 0.02 | 0.02 |
| *Codeine* | 0.13 | 0.13 | 0.13 | 0.13 | 0.14 | 0.14 | 0.14 | 0.14 | 0.14 |
| Systemic NSAIDs | 0.23 | 0.22 | 0.21 | 0.19 | 0.19 | 0.19 | 0.17 | 0.17 | 0.17 |
| *Non-selective NSAIDs* | 0.20 | 0.18 | 0.17 | 0.16 | 0.16 | 0.16 | 0.14 | 0.14 | 0.14 |
| *Coxib* | 0.03 | 0.03 | 0.03 | 0.03 | 0.03 | 0.03 | 0.03 | 0.03 | 0.03 |
| Paracetamol (incl. combinations) | 0.38 | 0.38 | 0.38 | 0.38 | 0.40 | 0.41 | 0.42 | 0.42 | 0.42 |
| *Paracetamol (excl. opioid combinations)* | 0.22 | 0.22 | 0.22 | 0.22 | 0.23 | 0.24 | 0.25 | 0.25 | 0.25 |
| Gabapentin | 0.02 | 0.02 | 0.02 | 0.02 | 0.02 | 0.02 | 0.02 | 0.02 | 0.02 |
| Pregabalin | 0.09 | 0.10 | 0.10 | 0.10 | 0.10 | 0.10 | 0.10 | 0.09 | 0.09 |
| Amitriptyline | 0.01 | 0.01 | 0.02 | 0.02 | 0.02 | 0.02 | 0.03 | 0.03 | 0.03 |
| Topical analgesics | 0.13 | 0.14 | 0.15 | 0.15 | 0.13 | 0.13 | 0.13 | 0.14 | 0.14 |
| *Topical NSAIDs* | 0.11 | 0.11 | 0.11 | 0.11 | 0.12 | 0.12 | 0.13 | 0.13 | 0.13 |
| *Lidocaine* | 0.02 | 0.03 | 0.04 | 0.04 | <0.01 | <0.01 | <0.01 | <0.01 | <0.01 |
| *Capsaicin* | <0.01 | <0.01 | <0.01 | <0.01 | <0.01 | <0.01 | <0.01 | <0.01 | <0.01 |
| Antimigraines | 0.01 | 0.01 | 0.01 | 0.01 | 0.01 | 0.01 | 0.01 | 0.01 | 0.01 |

| **Proportional share of opioid prescribing (Ireland)** | | | | | | | | | |
| --- | --- | --- | --- | --- | --- | --- | --- | --- | --- |
|  | **2014** | **2015** | **2016** | **2017** | **2018** | **2019** | **2020** | **2021** | **2022** |
| Strong opioids | 0.51 | 0.52 | 0.53 | 0.53 | 0.53 | 0.52 | 0.51 | 0.51 | 0.50 |
| Long-acting opioids | 0.21 | 0.22 | 0.22 | 0.23 | 0.23 | 0.23 | 0.23 | 0.22 | 0.22 |
| Morphine | 0.03 | 0.03 | 0.03 | 0.03 | 0.03 | 0.03 | 0.03 | 0.03 | 0.03 |
| Hydromorphone | <0.01 | <0.01 | <0.01 | <0.01 | <0.01 | <0.01 | <0.01 | <0.01 | <0.01 |
| Oxycodone | 0.08 | 0.08 | 0.09 | 0.09 | 0.09 | 0.09 | 0.09 | 0.10 | 0.09 |
| Dihydrocodeine | 0.04 | 0.01 | <0.01 | <0.01 | <0.01 | <0.01 | <0.01 | <0.01 | <0.01 |
| Pethidine | <0.01 | <0.01 | <0.01 | <0.01 | <0.01 | <0.01 | 0 | 0 | 0 |
| Fentanyl | 0.04 | 0.03 | 0.03 | 0.03 | 0.03 | 0.03 | 0.03 | 0.03 | 0.03 |
| Buprenorphine | 0.07 | 0.07 | 0.08 | 0.08 | 0.08 | 0.08 | 0.08 | 0.08 | 0.08 |
| Tramadol | 0.28 | 0.28 | 0.27 | 0.25 | 0.24 | 0.23 | 0.22 | 0.22 | 0.21 |
| Meptazinol | <0.01 | <0.01 | <0.01 | <0.01 | <0.01 | <0.01 | <0.01 | <0.01 | <0.01 |
| Tapentadol | 0.01 | 0.02 | 0.03 | 0.04 | 0.05 | 0.05 | 0.05 | 0.06 | 0.06 |
| Codeine | 0.45 | 0.47 | 0.47 | 0.47 | 0.47 | 0.48 | 0.49 | 0.49 | 0.50 |

| **Proportional share of analgesic prescribing (England)** | | | | | | | | | |
| --- | --- | --- | --- | --- | --- | --- | --- | --- | --- |
|  | **2014** | **2015** | **2016** | **2017** | **2018** | **2019** | **2020** | **2021** | **2022** |
| Opioids | 0.40 | 0.39 | 0.39 | 0.39 | 0.39 | 0.40 | 0.40 | 0.40 | 0.39 |
| *Strong opioids* | 0.16 | 0.16 | 0.16 | 0.16 | 0.16 | 0.16 | 0.17 | 0.16 | 0.16 |
| *Long-acting opioids* | 0.07 | 0.07 | 0.08 | 0.08 | 0.08 | 0.08 | 0.08 | 0.08 | 0.08 |
| *Morphine* | 0.04 | 0.04 | 0.04 | 0.05 | 0.05 | 0.05 | 0.05 | 0.05 | 0.05 |
| *Hydromorphone* | <0.01 | <0.01 | <0.01 | <0.01 | <0.01 | <0.01 | <0.01 | <0.01 | <0.01 |
| *Oxycodone* | 0.01 | 0.01 | 0.02 | 0.02 | 0.02 | 0.02 | 0.02 | 0.02 | 0.02 |
| *Dihydrocodeine* | 0.04 | 0.04 | 0.04 | 0.04 | 0.03 | 0.03 | 0.03 | 0.03 | 0.03 |
| *Pethidine* | <0.01 | <0.01 | <0.01 | <0.01 | <0.01 | <0.01 | <0.01 | <0.01 | <0.01 |
| *Fentanyl* | 0.01 | 0.01 | 0.01 | 0.01 | 0.01 | 0.01 | 0.01 | 0.01 | 0.01 |
| *Buprenorphine* | 0.02 | 0.02 | 0.02 | 0.02 | 0.02 | 0.02 | 0.02 | 0.02 | 0.02 |
| *Tramadol* | 0.08 | 0.07 | 0.07 | 0.07 | 0.06 | 0.06 | 0.06 | 0.06 | 0.06 |
| *Meptazinol* | <0.01 | <0.01 | <0.01 | <0.01 | <0.01 | <0.01 | <0.01 | <0.01 | <0.01 |
| *Tapentadol* | <0.01 | <0.01 | <0.01 | <0.01 | <0.01 | <0.01 | <0.01 | <0.01 | <0.01 |
| *Codeine* | 0.19 | 0.19 | 0.19 | 0.19 | 0.20 | 0.20 | 0.20 | 0.20 | 0.20 |
| Systemic NSAIDs | 0.14 | 0.14 | 0.13 | 0.12 | 0.11 | 0.11 | 0.10 | 0.10 | 0.11 |
| *Non-selective NSAIDs* | 0.14 | 0.13 | 0.12 | 0.11 | 0.10 | 0.10 | 0.09 | 0.10 | 0.10 |
| *Coxib* | 0.01 | 0.01 | 0.01 | 0.01 | 0.01 | 0.01 | 0.01 | 0.01 | 0.01 |
| Paracetamol (incl. combinations) | 0.40 | 0.39 | 0.38 | 0.36 | 0.35 | 0.34 | 0.34 | 0.33 | 0.32 |
| *Paracetamol (excl. combinations)* | 0.22 | 0.22 | 0.21 | 0.19 | 0.18 | 0.17 | 0.17 | 0.17 | 0.16 |
| Gabapentin | 0.05 | 0.05 | 0.06 | 0.07 | 0.07 | 0.07 | 0.07 | 0.07 | 0.07 |
| Pregabalin | 0.04 | 0.05 | 0.05 | 0.06 | 0.07 | 0.07 | 0.08 | 0.08 | 0.08 |
| Amitriptyline | 0.07 | 0.07 | 0.07 | 0.08 | 0.08 | 0.09 | 0.10 | 0.10 | 0.11 |
| Topical analgesics | 0.06 | 0.06 | 0.07 | 0.07 | 0.06 | 0.06 | 0.04 | 0.04 | 0.05 |
| *Topical NSAIDs* | 0.05 | 0.06 | 0.06 | 0.06 | 0.06 | 0.06 | 0.04 | 0.04 | 0.04 |
| *Lidocaine* | <0.01 | <0.01 | <0.01 | <0.01 | <0.01 | <0.01 | <0.01 | <0.01 | <0.01 |
| *Capsaicin* | <0.01 | <0.01 | <0.01 | <0.01 | <0.01 | <0.01 | <0.01 | <0.01 | <0.01 |
| Antimigraines | 0.03 | 0.03 | 0.03 | 0.03 | 0.03 | 0.03 | 0.03 | 0.03 | 0.04 |

| **Proportional share of opioid prescribing (NHS)** | | | | | | | | | |
| --- | --- | --- | --- | --- | --- | --- | --- | --- | --- |
|  | **2014** | **2015** | **2016** | **2017** | **2018** | **2019** | **2020** | **2021** | **2022** |
| Strong opioids | 0.40 | 0.41 | 0.41 | 0.42 | 0.41 | 0.42 | 0.41 | 0.41 | 0.41 |
| Long-acting opioids | 0.18 | 0.19 | 0.20 | 0.20 | 0.20 | 0.21 | 0.20 | 0.20 | 0.20 |
| Morphine | 0.10 | 0.11 | 0.11 | 0.12 | 0.12 | 0.13 | 0.13 | 0.13 | 0.13 |
| Hydromorphone | <0.01 | <0.01 | <0.01 | <0.01 | <0.01 | <0.01 | <0.01 | <0.01 | <0.01 |
| Oxycodone | 0.03 | 0.04 | 0.04 | 0.04 | 0.04 | 0.05 | 0.05 | 0.05 | 0.05 |
| Dihydrocodeine | 0.11 | 0.10 | 0.10 | 0.09 | 0.09 | 0.08 | 0.08 | 0.08 | 0.07 |
| Pethidine | <0.01 | <0.01 | <0.01 | <0.01 | <0.01 | <0.01 | <0.01 | <0.01 | <0.01 |
| Fentanyl | 0.03 | 0.03 | 0.03 | 0.03 | 0.03 | 0.03 | 0.02 | 0.02 | 0.02 |
| Buprenorphine | 0.05 | 0.05 | 0.05 | 0.06 | 0.06 | 0.06 | 0.06 | 0.06 | 0.06 |
| Tramadol | 0.19 | 0.18 | 0.18 | 0.17 | 0.16 | 0.15 | 0.15 | 0.15 | 0.14 |
| Meptazinol | <0.01 | <0.01 | <0.01 | <0.01 | <0.01 | <0.01 | <0.01 | <0.01 | <0.01 |
| Tapentadol | <0.01 | <0.01 | <0.01 | <0.01 | <0.01 | <0.01 | <0.01 | <0.01 | <0.01 |
| Codeine | 0.48 | 0.49 | 0.49 | 0.49 | 0.50 | 0.50 | 0.50 | 0.51 | 0.51 |

**Supplementary table 6. Rate ratios for Ireland versus England 2014-2022**

|  | **Rate ratios for dispensings in Ireland versus England** | | | | | | | | | |
| --- | --- | --- | --- | --- | --- | --- | --- | --- | --- | --- |
|  |  | **2014** | **2015** | **2016** | **2017** | **2018** | **2019** | **2020** | **2021** | **2022** |
|  | All analgesics | 1.86 | 1.94 | 2.05 | 2.18 | 2.28 | 2.41 | 2.47 | 2.64 | 2.85 |
|  | Opioids | 1.36 | 1.40 | 1.48 | 1.56 | 1.68 | 1.77 | 1.88 | 1.92 | 2.09 |
|  | *Strong opioids* | 1.71 | 1.79 | 1.88 | 1.99 | 2.14 | 2.20 | 2.48 | 2.34 | 2.50 |
|  | *Long-acting opioids* | 1.54 | 1.59 | 1.68 | 1.78 | 1.91 | 1.96 | 2.27 | 2.09 | 2.24 |
|  | *Morphine* | 0.41 | 0.39 | 0.38 | 0.39 | 0.42 | 0.44 | 0.47 | 0.46 | 0.49 |
|  | *Hydromorphone* | 19.67 | 19.35 | 17.46 | 15.44 | 16.46 | 17.04 | 19.96 | 17.54 | 14.99 |
|  | *Oxycodone* | 3.19 | 3.22 | 3.32 | 3.44 | 3.53 | 3.64 | 3.59 | 3.91 | 3.80 |
|  | *Dihydrocodeine* | 0.50 | 0.14 | 0.01 | <0.01 | <0.01 | <0.01 | <0.01 | <0.01 | <0.01 |
|  | *Pethidine* | 0.24 | 0.33 | 0.39 | 0.30 | 0.31 | 0.13 | 0 | 0 | 0 |
|  | *Fentanyl* | 1.62 | 1.61 | 1.68 | 1.79 | 1.99 | 2.15 | 2.25 | 2.46 | 2.81 |
|  | *Buprenorphine* | 2.04 | 2.11 | 2.15 | 2.22 | 2.35 | 2.33 | 2.29 | 2.35 | 2.48 |
|  | *Tramadol* | 1.94 | 2.12 | 2.25 | 2.38 | 2.58 | 2.67 | 2.66 | 2.80 | 3.06 |
|  | *Meptazinol* | 1.71 | 1.70 | 1.72 | 2.22 | 2.13 | 2.32 | 2.25 | 2.15 | 2.43 |
|  | *Tapentadol* | 12.25 | 14.29 | 17.31 | 19.27 | 21.00 | 22.05 | 22.94 | 25.50 | 26.03 |
|  | *Codeine* | 1.26 | 1.35 | 1.43 | 1.49 | 1.60 | 1.70 | 1.72 | 1.86 | 2.05 |
|  | Systemic NSAIDs | 3.02 | 3.13 | 3.34 | 3.67 | 4.00 | 4.15 | 4.22 | 4.34 | 4.39 |
|  | *Non-selective NSAIDs* | 2.69 | 2.78 | 2.97 | 3.25 | 3.56 | 3.73 | 3.78 | 3.91 | 4.00 |
|  | *Coxib* | 9.88 | 10.56 | 10.79 | 11.33 | 11.28 | 10.34 | 9.83 | 9.64 | 8.95 |
|  | Paracetamol (incl. combinations) | 1.76 | 1.88 | 2.05 | 2.30 | 2.61 | 2.89 | 3.00 | 3.33 | 3.77 |
|  | *Paracetamol (excl. combinations)* | 1.80 | 1.96 | 2.16 | 2.51 | 2.97 | 3.38 | 3.53 | 3.98 | 4.57 |
|  | Gabapentin | 0.63 | 0.58 | 0.58 | 0.59 | 0.63 | 0.70 | 0.75 | 0.84 | 0.98 |
|  | Pregabalin | 4.34 | 4.14 | 3.97 | 3.71 | 3.41 | 3.23 | 2.98 | 2.94 | 3.12 |
|  | Amitriptyline | 0.35 | 0.39 | 0.45 | 0.52 | 0.58 | 0.64 | 0.67 | 0.70 | 0.76 |
|  | Topical analgesics | 4.10 | 4.38 | 4.67 | 5.03 | 4.42 | 5.28 | 7.48 | 8.21 | 8.34 |
|  | *Topical NSAIDs* | 3.57 | 3.61 | 3.66 | 4.04 | 4.57 | 5.38 | 7.83 | 8.73 | 8.79 |
|  | *Lidocaine* | 23.37 | 28.44 | 33.54 | 34.08 | 3.10 | 5.34 | 6.77 | 6.90 | 6.64 |
|  | *Capsaicin* | 1.50 | 1.56 | 1.41 | 1.64 | 2.15 | 2.52 | 1.29 | 1.63 | 1.98 |
|  | Antimigraines | 0.97 | 0.99 | 1.03 | 1.06 | 1.05 | 0.92 | 0.91 | 0.96 | 1.00 |
| **Rate ratios for costs (adjusted for exchange rate and purchasing power parity*)** \| \| \| \| \| \| \| \| \|; \| **2014** \| **2015** \| **2016** \| **2017** \| **2018** \| **2019** \| **2020** \| **2021** \| **2022**; All analgesics \| 3.98 \| 3.74 \| 4.48 \| 5.28 \| 6.08 \| 6.13 \| 5.70 \| 5.67 \| 6.52; Opioids \| 2.76 \| 2.55 \| 3.30 \| 3.94 \| 4.43 \| 4.58 \| 5.16 \| 4.58 \| 5.09; *Strong opioids* \| 3.09 \| 2.95 \| 3.66 \| 4.26 \| 4.61 \| 4.87 \| 6.42 \| 5.26 \| 5.35; *Longacting opioids* \| 2.52 \| 2.42 \| 3.01 \| 3.72 \| 4.07 \| 4.32 \| 5.65 \| 4.76 \| 4.66; *Morphine* \| 0.71 \| 0.63 \| 0.71 \| 0.76 \| 0.81 \| 0.88 \| 0.92 \| 0.87 \| 0.90; *Hydromorphone* \| 21.36 \| 21.24 \| 20.53 \| 21.61 \| 22.67 \| 25.80 \| 30.79 \| 26.84 \| 24.33; *Oxycodone* \| 2.13 \| 1.96 \| 2.52 \| 3.12 \| 3.50 \| 4.01 \| 4.45 \| 4.72 \| 4.53; *Dihydrocodeine* \| 0.59 \| 0.15 \| 0.01 \| <0.01 \| <0.01 \| <0.01 \| <0.01 \| <0.01 \| <0.01; *Pethidine* \| 0.02 \| 0.03 \| 0.05 \| 0.05 \| 0.07 \| 0.02 \| 0 \| 0 \| 0; *Fentanyl* \| 3.50 \| 3.24 \| 3.73 \| 4.01 \| 4.40 \| 4.59 \| 4.90 \| 5.12 \| 5.45; *Buprenorphine* \| 3.71 \| 3.41 \| 3.90 \| 5.10 \| 5.20 \| 4.76 \| 4.64 \| 4.62 \| 4.05; *Tramadol* \| 3.97 \| 4.00 \| 5.57 \| 5.71 \| 6.29 \| 6.91 \| 6.75 \| 6.76 \| 8.13; *Meptazinol* \| 2.19 \| 2.04 \| 2.30 \| 3.06 \| 2.58 \| 2.82 \| 2.82 \| 2.65 \| 2.90; *Tapentadol* \| 13.15 \| 12.93 \| 16.90 \| 20.42 \| 21.95 \| 23.46 \| 24.93 \| 26.20 \| 25.72; *Codeine* \| 2.35 \| 2.07 \| 2.96 \| 3.80 \| 4.66 \| 4.61 \| 4.15 \| 4.20 \| 5.85; Systemic NSAIDs \| 6.16 \| 5.89 \| 7.85 \| 8.25 \| 7.72 \| 3.87 \| 6.65 \| 8.62 \| 10.77; *Non-selective NSAIDs* \| 4.80 \| 4.27 \| 5.75 \| 6.08 \| 6.07 \| 2.99 \| 5.18 \| 7.21 \| 9.80; *Coxib* \| 10.62 \| 13.93 \| 18.39 \| 20.24 \| 25.77 \| 24.44 \| 33.73 \| 21.27 \| 16.18; Paracetamol (incl. combinations) \| 2.92 \| 2.65 \| 3.67 \| 4.58 \| 6.02 \| 5.87 \| 4.91 \| 4.99 \| 6.49; *Paracetamol (excl. combinations)* \| 2.11 \| 2.00 \| 2.84 \| 3.95 \| 6.27 \| 5.79 \| 4.47 \| 4.62 \| 5.93; Gabapentin \| 2.55 \| 2.71 \| 3.45 \| 2.52 \| 2.78 \| 4.38 \| 3.16 \| 3.32 \| 3.84; Pregabalin \| 4.10 \| 3.30 \| 3.05 \| 4.45 \| 25.22 \| 34.31 \| 19.36 \| 9.78 \| 16.06; Amitriptyline \| 1.33 \| 1.27 \| 1.71 \| 2.20 \| 2.27 \| 3.07 \| 2.63 \| 2.79 \| 3.85; Topical analgesics \| 15.74 \| 17.60 \| 21.96 \| 24.62 \| 9.74 \| 13.23 \| 18.86 \| 19.18 \| 18.98; *Topical NSAIDs* \| 7.03 \| 6.90 \| 8.72 \| 11.41 \| 13.71 \| 16.80 \| 27.82 \| 29.72 \| 29.05; *Lidocaine* \| 39.95 \| 41.75 \| 49.71 \| 52.14 \| 5.40 \| 9.62 \| 12.37 \| 11.93 \| 10.83; *Capsaicin* \| 2.22 \| 2.13 \| 2.12 \| 2.64 \| 3.51 \| 4.25 \| 2.24 \| 2.67 \| 3.12; Antimigraines \| 2.28 \| 2.04 \| 2.20 \| 1.00 \| 1.36 \| 1.83 \| 1.97 \| 2.14 \| 1.96 *The World Bank. Word Development Indicators. <https://databank.worldbank.org/reports.aspx?source=2&series=PA.NUS.FCRF&country>= | | | | | | | | | | |

| **Rate ratios for DDDs in Ireland versus England** | | | | | | | | | |
| --- | --- | --- | --- | --- | --- | --- | --- | --- | --- |
|  | **2014** | **2015** | **2016** | **2017** | **2018** | **2019** | **2020** | **2021** | **2022** |
| All analgesics | 1.20 | 1.25 | 1.31 | 1.39 | 1.49 | 1.57 | 1.60 | 1.71 | 1.82 |
| Opioids | 0.84 | 0.87 | 0.91 | 0.95 | 1.01 | 1.07 | 1.09 | 1.17 | 1.26 |
| *Strong opioids* | 1.25 | 1.32 | 1.41 | 1.51 | 1.65 | 1.75 | 1.81 | 1.94 | 2.11 |
| *Long-acting opioids* | 1.12 | 1.17 | 1.24 | 1.33 | 1.45 | 1.55 | 1.62 | 1.74 | 1.92 |
| *Morphine* | 0.23 | 0.23 | 0.23 | 0.24 | 0.25 | 0.27 | 0.30 | 0.30 | 0.32 |
| *Hydromorphone* | 22.87 | 24.83 | 21.41 | 22.15 | 21.54 | 24.49 | 28.70 | 24.64 | 23.36 |
| *Oxycodone* | 1.92 | 2.00 | 2.12 | 2.23 | 2.36 | 2.53 | 2.64 | 2.91 | 2.91 |
| *Dihydrocodeine* | 0.29 | 0.11 | 0.01 | <0.01 | <0.01 | <0.01 | <0.01 | <0.01 | <0.01 |
| *Pethidine* | 0.04 | 0.06 | 0.12 | 0.14 | 0.16 | 0.07 | 0 | 0 | 0 |
| *Fentanyl* | 1.85 | 1.64 | 1.70 | 1.79 | 1.96 | 2.15 | 2.27 | 2.50 | 2.86 |
| *Buprenorphine* | 1.66 | 1.71 | 1.75 | 1.83 | 1.94 | 1.99 | 2.01 | 2.09 | 2.24 |
| *Tramadol* | 1.27 | 1.40 | 1.51 | 1.61 | 1.76 | 1.85 | 1.88 | 1.99 | 2.16 |
| *Meptazinol* | 1.33 | 1.37 | 1.39 | 1.72 | 1.61 | 1.73 | 1.71 | 1.70 | 1.97 |
| *Tapentadol* | 9.40 | 10.25 | 12.03 | 13.49 | 14.46 | 15.16 | 15.86 | 17.40 | 17.85 |
| *Codeine* | 0.71 | 0.74 | 0.78 | 0.80 | 0.84 | 0.89 | 0.91 | 0.98 | 1.06 |
| Systemic NSAIDs | 1.93 | 2.03 | 2.17 | 2.38 | 2.58 | 2.69 | 2.76 | 2.81 | 2.78 |
| *Non-selective NSAIDs* | 1.63 | 1.71 | 1.83 | 2.01 | 2.19 | 2.30 | 2.37 | 2.42 | 2.42 |
| *Coxib* | 7.56 | 8.15 | 8.38 | 8.90 | 9.02 | 8.40 | 8.09 | 8.03 | 7.37 |
| Paracetamol (incl. combinations) | 2.41 | 2.60 | 2.85 | 3.26 | 3.74 | 4.19 | 4.33 | 4.82 | 5.43 |
| *Paracetamol (excl. combinations)* | 1.61 | 1.77 | 1.95 | 2.26 | 2.62 | 2.94 | 3.08 | 3.43 | 3.87 |
| Gabapentin | 0.48 | 0.45 | 0.44 | 0.44 | 0.46 | 0.51 | 0.54 | 0.60 | 0.70 |
| Pregabalin | 2.96 | 2.90 | 2.84 | 2.70 | 2.53 | 2.41 | 2.23 | 2.22 | 2.35 |
| Amitriptyline | 0.24 | 0.27 | 0.32 | 0.38 | 0.42 | 0.46 | 0.49 | 0.52 | 0.55 |
| Antimigraines | 0.85 | 0.89 | 0.94 | 1.00 | 0.95 | 0.51 | 0.63 | 0.65 | 0.67 |

| **Rate ratios for OMEs in Ireland versus England** | | | | | | | | | |
| --- | --- | --- | --- | --- | --- | --- | --- | --- | --- |
|  | **2014** | **2015** | **2016** | **2017** | **2018** | **2019** | **2020** | **2021** | **2022** |
| Opioids | 1.12 | 1.19 | 1.27 | 1.37 | 1.49 | 1.58 | 1.64 | 1.76 | 1.91 |
| *Strong opioids* | 1.25 | 1.35 | 1.45 | 1.57 | 1.73 | 1.85 | 1.93 | 2.08 | 2.26 |
| *Long-acting opioids* | 1.25 | 1.31 | 1.41 | 1.53 | 1.67 | 1.80 | 1.89 | 2.05 | 2.25 |
| *Morphine* | 0.23 | 0.23 | 0.23 | 0.23 | 0.25 | 0.27 | 0.29 | 0.30 | 0.32 |
| *Hydromorphone* | 17.22 | 19.82 | 17.09 | 16.19 | 18.02 | 20.72 | 25.08 | 22.78 | 22.11 |
| *Oxycodone* | 1.90 | 1.98 | 2.10 | 2.21 | 2.34 | 2.51 | 2.62 | 2.89 | 2.89 |
| *Dihydrocodeine* | 0.29 | 0.11 | 0.01 | <0.01 | <0.01 | <0.01 | <0.01 | <0.01 | <0.01 |
| *Pethidine* | 0.14 | 0.22 | 0.41 | 0.48 | 0.56 | 0.22 | 0 | 0 | 0 |
| *Fentanyl* | 1.55 | 1.57 | 1.63 | 1.74 | 1.91 | 2.10 | 2.23 | 2.44 | 2.81 |
| *Buprenorphine* | 1.86 | 1.90 | 1.93 | 2.00 | 2.11 | 2.13 | 2.14 | 2.22 | 2.36 |
| *Tramadol* | 1.27 | 1.40 | 1.51 | 1.61 | 1.76 | 1.85 | 1.88 | 1.99 | 2.16 |
| *Meptazinol* | 1.33 | 1.37 | 1.39 | 1.72 | 1.61 | 1.73 | 1.71 | 1.70 | 1.97 |
| *Tapentadol* | 9.40 | 10.25 | 12.03 | 13.49 | 14.46 | 15.16 | 15.86 | 17.40 | 17.85 |
| *Codeine* | 0.71 | 0.74 | 0.78 | 0.80 | 0.84 | 0.89 | 0.91 | 0.98 | 1.06 |

**Supplementary table 7. Cost rates for Ireland and England**

| **Rate of costs (in €) per 1,000 GMS population** | | | | | | | | | | | |
| --- | --- | --- | --- | --- | --- | --- | --- | --- | --- | --- | --- |
|  | **2014** | **2015** | **2016** | **2017** | **2018** | **2019** | **2020** | **2021** | **2022** | **Absolute change** | **Relative change** |
| All analgesics | 63972.85 | 70620.13 | 73052.30 | 71586.05 | 58276.80 | 58963.54 | 51917.20 | 53197.13 | 51988.38 | -11984.47* | -0.19 |
| Opioids | 18892.92 | 20158.43 | 21178.51 | 20757.96 | 20956.56 | 20831.33 | 20784.10 | 21792.21 | 20663.43 | 1770.51 | 0.09 |
| *Strong opioids* | 14432.75 | 15435.41 | 16173.66 | 15531.23 | 15285.82 | 14837.79 | 14574.84 | 15048.36 | 13897.20 | -535.55 | -0.04 |
| *Long-acting opioids* | 8773.30 | 9423.19 | 10015.81 | 10003.99 | 9992.00 | 9704.71 | 9398.08 | 9592.64 | 8701.97 | -71.33 | -0.01 |
| *Morphine* | 395.35 | 409.81 | 424.76 | 417.99 | 426.26 | 447.76 | 466.90 | 432.77 | 402.35 | 7.00 | 0.02 |
| *Hydromorphone* | 153.29 | 160.80 | 129.44 | 115.69 | 105.47 | 109.59 | 118.12 | 91.17 | 73.33 | -79.97* | -0.52 |
| *Oxycodone* | 2345.97 | 2375.25 | 2428.83 | 2559.06 | 2706.54 | 2808.24 | 2805.37 | 2928.99 | 2705.69 | 359.72* | 0.15 |
| *Dihydrocodeine* | 213.19 | 64.68 | 3.05 | 0.65 | 0.27 | 0.19 | 0.10 | 0.04 | 0.03 | -213.16* | -1.00 |
| *Pethidine* | 0.82 | 1.14 | 1.50 | 1.32 | 1.50 | 0.41 | 0 | 0 | 0 | 0 | 0 |
| *Fentanyl* | 3437.93 | 3471.49 | 3487.75 | 3197.61 | 3170.26 | 2840.27 | 2707.38 | 2680.78 | 2478.84 | -959.09* | -0.28 |
| *Buprenorphine* | 3759.04 | 4095.11 | 4341.01 | 4175.06 | 3862.65 | 3381.54 | 3095.47 | 3179.85 | 2646.51 | -1112.53* | -0.30 |
| *Tramadol* | 3647.97 | 3885.13 | 3796.48 | 3022.12 | 2627.02 | 2594.84 | 2558.59 | 2699.65 | 2590.90 | -1057.06* | -0.29 |
| *Meptazinol* | 43.07 | 44.30 | 44.22 | 53.27 | 44.14 | 46.27 | 43.81 | 41.67 | 41.60 | -1.47 | -0.03 |
| *Tapentadol* | 692.39 | 1036.68 | 1563.90 | 2042.39 | 2386.13 | 2655.14 | 2823.02 | 3035.15 | 2999.58 | 2307.19* | 3.33 |
| *Codeine* | 4203.92 | 4614.05 | 4957.59 | 5172.80 | 5626.32 | 5947.08 | 6165.35 | 6702.14 | 6724.61 | 2520.69* | 0.60 |
| Systemic NSAIDs | 7664.54 | 7821.89 | 7848.83 | 7712.71 | 7078.60 | 6987.44 | 6840.05 | 7383.83 | 7130.87 | -533.67 | -0.07 |
| *Non-selective NSAIDs* | 4564.30 | 4718.97 | 4791.72 | 4808.57 | 5092.87 | 5166.78 | 5057.66 | 5560.23 | 5509.80 | 945.50* | 0.21 |
| *Coxib* | 3100.24 | 3102.92 | 3057.11 | 2904.13 | 1985.74 | 1820.66 | 1782.40 | 1823.60 | 1621.07 | -1479.17* | -0.48 |
| Paracetamol (incl. combinations) | 9401.95 | 10106.38 | 10468.18 | 10343.18 | 10897.47 | 11606.99 | 12184.84 | 13396.87 | 13679.16 | 4277.21* | 0.45 |
| *Paracetamol (excl. combinations)* | 3130.24 | 3365.69 | 3535.24 | 3837.63 | 4281.61 | 4622.41 | 4935.59 | 5494.22 | 5733.28 | 2603.04* | 0.83 |
| Gabapentin | 1583.79 | 1633.72 | 1752.28 | 1817.40 | 1991.58 | 2162.76 | 1819.42 | 1813.48 | 1925.62 | 341.83* | 0.22 |
| Pregabalin | 17448.10 | 17672.53 | 16145.86 | 14820.78 | 14972.31 | 14698.44 | 7604.32 | 5638.64 | 5577.27 | -11870.83* | -0.68 |
| Amitriptyline | 270.51 | 339.32 | 409.97 | 493.63 | 570.26 | 652.12 | 714.33 | 812.71 | 880.02 | 609.51* | 2.25 |
| Topical analgesics | 13655.62 | 18196.02 | 20783.03 | 20898.45 | 7121.15 | 7688.23 | 7861.14 | 8776.80 | 8618.25 | -5037.38 | -0.37 |
| *Topical NSAIDs* | 3805.21 | 4211.86 | 4615.59 | 5277.51 | 5527.06 | 5669.36 | 5927.90 | 6837.23 | 6800.20 | 2994.99* | 0.79 |
| *Lidocaine* | 9662.70 | 13786.05 | 15963.50 | 15380.10 | 1266.97 | 1762.31 | 1803.64 | 1716.30 | 1589.25 | -8073.45 | -0.84 |
| *Capsaicin* | 187.72 | 198.11 | 203.94 | 240.84 | 327.13 | 256.56 | 129.61 | 223.28 | 228.80 | 41.08 | 0.22 |
| Antimigraines | 1327.13 | 1432.54 | 1398.57 | 1247.48 | 1305.39 | 1339.76 | 1382.42 | 1511.71 | 1487.74 | 160.61 | 0.12 |

| **Rate of costs (in £) per 1,000 NHS population** | | | | | | | | | | | |
| --- | --- | --- | --- | --- | --- | --- | --- | --- | --- | --- | --- |
|  | **2014** | **2015** | **2016** | **2017** | **2018** | **2019** | **2020** | **2021** | **2022** | **Absolute change** | **Relative change** |
| All analgesics | 15234.16 | 16013.91 | 15411.94 | 13798.16 | 9767.52 | 10008.95 | 9630.66 | 9453.87 | 7722.79 | -7511.37* | -0.49 |
| Opioids | 6472.38 | 6705.12 | 6077.45 | 5365.29 | 4819.77 | 4730.24 | 4254.94 | 4794.72 | 3931.60 | -2540.78* | -0.39 |
| *Strong opioids* | 4416.91 | 4441.97 | 4180.78 | 3715.29 | 3379.89 | 3167.36 | 2398.49 | 2880.54 | 2515.74 | -1901.17* | -0.43 |
| *Long-acting opioids* | 3296.45 | 3308.14 | 3145.84 | 2740.58 | 2501.90 | 2337.10 | 1758.22 | 2029.96 | 1805.71 | -1490.74* | -0.45 |
| *Morphine* | 527.05 | 553.32 | 568.59 | 561.88 | 534.19 | 527.99 | 534.47 | 503.43 | 432.69 | -94.35 | -0.18 |
| *Hydromorphine* | 6.79 | 6.43 | 5.96 | 5.45 | 4.74 | 4.42 | 4.06 | 3.42 | 2.92 | -3.88* | -0.57 |
| *Oxycodone* | 1043.82 | 1028.11 | 910.45 | 834.77 | 787.78 | 727.60 | 666.62 | 624.69 | 577.61 | -466.21* | -0.45 |
| *Dihydrocodeine* | 344.19 | 354.79 | 293.38 | 247.42 | 191.58 | 204.87 | 269.41 | 291.25 | 289.34 | -54.85 | -0.16 |
| *Pethidine* | 32.82 | 32.32 | 29.18 | 25.44 | 23.01 | 17.60 | 10.70 | 8.49 | 8.84 | -23.99* | -0.73 |
| *Fentanyl* | 929.19 | 909.03 | 883.15 | 813.11 | 735.34 | 643.38 | 584.31 | 527.78 | 440.32 | -488.87* | -0.53 |
| *Buprenorphine* | 958.23 | 1019.50 | 1051.62 | 833.43 | 758.13 | 738.35 | 705.87 | 693.65 | 631.96 | -326.27* | -0.34 |
| *Tramadol* | 869.16 | 825.17 | 644.31 | 539.32 | 425.83 | 390.33 | 400.48 | 402.32 | 308.53 | -560.63* | -0.65 |
| *Meptazinol* | 18.60 | 18.48 | 18.22 | 17.73 | 17.44 | 17.07 | 16.42 | 15.83 | 13.91 | -4.70* | -0.25 |
| *Tapentadol* | 49.85 | 68.10 | 87.53 | 101.89 | 110.87 | 117.69 | 119.71 | 116.75 | 112.88 | 63.03* | 1.26 |
| *Codeine* | 1693.17 | 1890.32 | 1585.48 | 1385.19 | 1231.12 | 1341.25 | 1570.91 | 1607.37 | 1112.87 | -580.30 | -0.34 |
| Systemic NSAIDs | 1177.64 | 1128.23 | 945.52 | 952.00 | 934.51 | 1876.95 | 1087.72 | 863.08 | 640.92 | -536.72 | -0.46 |
| *Non-selective NSAIDs* | 901.20 | 939.11 | 788.31 | 805.86 | 855.95 | 1799.48 | 1031.84 | 776.67 | 543.95 | -357.25 | -0.40 |
| *Coxib* | 276.44 | 189.12 | 157.20 | 146.14 | 78.57 | 77.47 | 55.87 | 86.41 | 96.97 | -179.47* | -0.65 |
| Paracetamol (incl. combinations) | 3045.87 | 3243.33 | 2698.95 | 2297.81 | 1845.67 | 2056.03 | 2621.10 | 2705.88 | 2040.68 | -1005.18 | -0.33 |
| *Paracetamol (excl. combinations)* | 1402.86 | 1425.27 | 1178.44 | 990.06 | 696.62 | 829.93 | 1168.16 | 1198.10 | 935.15 | -467.71 | -0.33 |
| Gabapentin | 588.22 | 512.18 | 480.17 | 735.73 | 729.62 | 513.71 | 608.09 | 550.41 | 485.12 | -103.11 | -0.18 |
| Pregabalin | 4029.20 | 4543.86 | 5007.54 | 3394.92 | 605.29 | 445.57 | 415.22 | 581.19 | 336.11 | -3693.09* | -0.92 |
| Amitriptyline | 191.84 | 226.45 | 226.22 | 228.35 | 255.67 | 221.12 | 287.14 | 293.44 | 221.12 | 29.27 | 0.15 |
| Topical analgesics | 821.49 | 877.71 | 894.94 | 864.43 | 745.64 | 604.30 | 440.62 | 461.10 | 439.62 | -381.87* | -0.46 |
| *Topical NSAIDs* | 512.48 | 518.38 | 500.38 | 470.94 | 411.20 | 350.96 | 225.30 | 231.79 | 226.56 | -285.92* | -0.56 |
| *Lidocaine* | 229.01 | 280.38 | 303.68 | 300.47 | 239.29 | 190.48 | 154.14 | 145.02 | 142.01 | -87.00* | -0.38 |
| *Capsaicin* | 80.00 | 78.96 | 90.87 | 93.02 | 95.14 | 62.86 | 61.18 | 84.29 | 71.05 | -8.95 | -0.11 |
| Antimigraines | 550.52 | 595.09 | 601.67 | 1267.38 | 980.39 | 760.85 | 741.04 | 711.84 | 733.15 | 182.63 | 0.33 |

**Supplemental table 8. Rate of costs, DDDs, and OMEs per 1,000 NHS population per day and percentage share of analgesics and opioids (top 33% of CCGs for deprivation)**

**Rate of costs**

|  | **2014** | **2015** | **2016** | **2017** | **2018** | **2019** | **2020** | **2021** | **2022** | **Absolute change** | **Relative change** |
| --- | --- | --- | --- | --- | --- | --- | --- | --- | --- | --- | --- |
| All analgesics | 16897.21 | 18050.59 | 17280.98 | 15096.31 | 10647.11 | 10991.45 | 10914.58 | 10833.69 | 9367.62 | -7529.59 | -0.45 |
| Opioids | 7297.93 | 7674.90 | 6897.08 | 5969.45 | 5341.97 | 5335.63 | 5665.69 | 5629.76 | 4860.13 | -2437.80 | -0.33 |
| *Strong opioids* | 4875.76 | 4961.23 | 4627.25 | 4017.71 | 3638.17 | 3452.01 | 3344.79 | 3219.68 | 2961.35 | -1914.41 | -0.39 |
| *Long-acting opioids* | 3699.56 | 3774.85 | 3567.21 | 3039.84 | 2761.77 | 2610.76 | 2437.93 | 2294.45 | 2166.55 | -1533.01 | -0.41 |
| *Morphine* | 571.58 | 613.47 | 635.11 | 624.31 | 589.14 | 583.22 | 596.78 | 566.31 | 533.59 | -37.99 | -0.07 |
| *Hydromorphone* | 7.86 | 7.20 | 7.05 | 6.25 | 5.86 | 5.37 | 5.53 | 4.16 | 4.17 | -3.68 | -0.47 |
| *Oxycodone* | 1170.30 | 1167.05 | 1038.46 | 921.93 | 875.34 | 813.00 | 759.47 | 710.95 | 698.63 | -471.67 | -0.40 |
| *Dihydrocodeine* | 389.05 | 411.67 | 343.88 | 288.15 | 227.82 | 245.62 | 331.96 | 350.74 | 363.07 | -25.99 | -0.07 |
| *Pethidine* | 28.27 | 28.92 | 26.63 | 21.28 | 18.96 | 14.20 | 6.93 | 5.57 | 6.34 | -21.93 | -0.78 |
| *Fentanyl* | 974.58 | 962.23 | 925.97 | 817.04 | 745.10 | 664.23 | 615.90 | 566.30 | 493.56 | -481.02 | -0.49 |
| *Buprenorphine* | 981.02 | 1053.15 | 1095.77 | 856.02 | 767.75 | 766.13 | 721.11 | 723.34 | 675.41 | -305.62 | -0.31 |
| *Tramadol* | 1098.04 | 1063.39 | 810.40 | 667.01 | 521.56 | 485.07 | 510.47 | 518.10 | 434.80 | -663.24 | -0.60 |
| *Meptazinol* | 19.07 | 18.30 | 17.15 | 15.54 | 14.15 | 12.95 | 11.32 | 9.11 | 8.44 | -10.63 | -0.56 |
| *Tapentadol* | 44.10 | 65.83 | 87.86 | 103.86 | 114.46 | 120.80 | 128.61 | 124.95 | 114.86 | 70.75 | 1.60 |
| *Codeine* | 2014.37 | 2283.95 | 1909.03 | 1648.25 | 1462.01 | 1625.25 | 1977.77 | 2050.35 | 1527.39 | -486.98 | -0.24 |
| Systemic NSAIDs | 1228.38 | 1198.01 | 994.58 | 1005.39 | 980.39 | 1949.16 | 1149.76 | 936.17 | 737.88 | -490.50 | -0.40 |
| *Non-selective NSAIDs* | 952.24 | 1010.81 | 843.07 | 866.70 | 906.40 | 1873.30 | 1093.95 | 850.81 | 628.30 | -323.94 | -0.34 |
| *Coxib* | 276.14 | 187.19 | 151.52 | 138.69 | 74.00 | 75.86 | 55.81 | 85.36 | 109.58 | -166.56 | -0.60 |
| Paracetamol (incl. opioid combinations) | 3556.34 | 3856.20 | 3197.47 | 2679.94 | 2146.95 | 2418.01 | 3165.10 | 3279.02 | 2633.02 | -923.32 | -0.26 |
| *Paracetamol (excl. opioid combinations)* | 1615.09 | 1671.87 | 1377.08 | 1129.79 | 784.06 | 932.49 | 1357.18 | 1384.82 | 1167.52 | -447.57 | -0.28 |
| Gabapentin | 678.61 | 604.30 | 565.78 | 859.46 | 855.25 | 611.12 | 729.35 | 655.80 | 612.35 | -66.26 | -0.10 |
| Pregabalin | 4464.78 | 5102.84 | 5617.80 | 3746.42 | 663.61 | 496.07 | 479.94 | 676.62 | 416.69 | -4048.09 | -0.91 |
| Amitriptyline | 179.18 | 215.56 | 215.48 | 215.27 | 242.18 | 237.18 | 279.59 | 285.20 | 236.11 | 56.93 | 0.32 |
| Topical analgesics | 963.06 | 1047.29 | 1072.57 | 1046.61 | 917.60 | 753.17 | 565.72 | 597.74 | 607.00 | -356.06 | -0.37 |
| *Topical NSAIDs* | 607.53 | 630.79 | 612.50 | 579.13 | 515.35 | 447.44 | 291.61 | 307.93 | 316.98 | -290.55 | -0.48 |
| *Lidocaine* | 259.12 | 318.38 | 345.07 | 349.66 | 280.15 | 223.26 | 198.46 | 183.45 | 188.42 | -70.70 | -0.27 |
| *Capsaicin* | 96.41 | 98.11 | 115.01 | 117.82 | 122.11 | 82.46 | 75.66 | 106.35 | 101.60 | 5.19 | 0.05 |
| Antimigraines | 470.17 | 535.82 | 540.61 | 1123.93 | 862.04 | 676.63 | 687.35 | 667.57 | 729.93 | 259.76 | 0.55 |

**Rate of DDDs**

|  | **2014** | **2015** | **2016** | **2017** | **2018** | **2019** | **2020** | **2021** | **2022** | **Absolute change** | **Relative change** |
| --- | --- | --- | --- | --- | --- | --- | --- | --- | --- | --- | --- |
| All analgesics | 108.21 | 110.98 | 110.37 | 105.37 | 102.10 | 100.43 | 101.56 | 101.18 | 102.01 | -6.19 | -0.06 |
| Opioids | 45.94 | 46.93 | 46.93 | 45.45 | 44.30 | 43.99 | 44.30 | 43.15 | 42.89 | -3.04 | -0.07 |
| *Strong opioids* | 13.89 | 13.77 | 13.55 | 12.76 | 11.85 | 11.40 | 11.04 | 10.56 | 10.17 | -3.73 | -0.27 |
| *Long-acting opioids* | 6.86 | 6.99 | 6.97 | 6.64 | 6.27 | 6.04 | 5.76 | 5.37 | 5.17 | -1.70 | -0.25 |
| *Morphine* | 2.42 | 2.57 | 2.66 | 2.60 | 2.47 | 2.43 | 2.32 | 2.19 | 2.11 | -0.30 | -0.13 |
| *Hydromorphone* | 0.01 | 0.01 | 0.01 | 0.01 | 0.01 | 0.01 | 0.01 | 0.00 | 0.00 | 0.00 | -0.48 |
| *Oxycodone* | 1.01 | 1.05 | 1.09 | 1.09 | 1.07 | 1.04 | 1.00 | 0.94 | 0.96 | -0.05 | -0.05 |
| *Dihydrocodeine* | 3.44 | 3.34 | 3.16 | 2.91 | 2.76 | 2.60 | 2.59 | 2.39 | 2.29 | -1.15 | -0.33 |
| *Pethidine* | 0.01 | 0.01 | 0.01 | 0.01 | 0.01 | 0.01 | 0.00 | 0.00 | 0.00 | -0.01 | -0.82 |
| *Fentanyl* | 1.34 | 1.36 | 1.34 | 1.26 | 1.16 | 1.06 | 0.97 | 0.87 | 0.79 | -0.55 | -0.41 |
| *Buprenorphine* | 0.70 | 0.73 | 0.76 | 0.77 | 0.76 | 0.77 | 0.77 | 0.76 | 0.74 | 0.04 | 0.06 |
| *Tramadol* | 8.37 | 7.98 | 7.61 | 6.95 | 6.28 | 5.98 | 5.88 | 5.69 | 5.46 | -2.91 | -0.35 |
| *Meptazinol* | 0.05 | 0.05 | 0.04 | 0.04 | 0.04 | 0.03 | 0.03 | 0.02 | 0.02 | -0.03 | -0.56 |
| *Tapentadol* | 0.04 | 0.05 | 0.07 | 0.09 | 0.09 | 0.10 | 0.11 | 0.10 | 0.09 | 0.06 | 1.58 |
| *Codeine* | 28.56 | 29.79 | 30.18 | 29.74 | 29.66 | 29.96 | 30.64 | 30.18 | 30.42 | 1.86 | 0.07 |
| Systemic NSAIDs | 25.07 | 24.80 | 23.51 | 21.15 | 19.70 | 19.00 | 18.09 | 19.22 | 20.47 | -4.60 | -0.18 |
| *Non-selective NSAIDs* | 23.83 | 23.60 | 22.33 | 20.05 | 18.61 | 17.86 | 16.86 | 17.91 | 19.01 | -4.82 | -0.20 |
| *Coxib* | 1.24 | 1.20 | 1.18 | 1.10 | 1.08 | 1.15 | 1.23 | 1.31 | 1.46 | 0.22 | 0.18 |
| Paracetamol (incl. opioid combinations) | 24.64 | 24.85 | 24.04 | 21.82 | 20.21 | 19.11 | 20.09 | 19.15 | 18.69 | -5.94 | -0.24 |
| *Paracetamol (excl. opioid combinations)* | 23.78 | 23.97 | 23.17 | 20.97 | 19.36 | 18.26 | 19.24 | 18.31 | 17.86 | -5.93 | -0.25 |
| Gabapentin | 4.55 | 5.30 | 5.90 | 6.35 | 6.55 | 6.37 | 6.47 | 6.37 | 6.28 | 1.73 | 0.38 |
| Pregabalin | 5.01 | 5.86 | 6.57 | 7.10 | 7.73 | 8.13 | 8.78 | 9.17 | 9.05 | 4.04 | 0.81 |
| Amitriptyline | 2.12 | 2.31 | 2.44 | 2.51 | 2.60 | 2.73 | 2.91 | 3.05 | 3.43 | 1.31 | 0.62 |
| Antimigraines | 1.74 | 1.82 | 1.84 | 1.84 | 1.86 | 1.94 | 1.77 | 1.90 | 2.03 | 0.29 | 0.17 |

**Rate of OMEs**

|  | **2014** | **2015** | **2016** | **2017** | **2018** | **2019** | **2020** | **2021** | **2022** | **Absolute change** | **Relative change** |
| --- | --- | --- | --- | --- | --- | --- | --- | --- | --- | --- | --- |
| All opioids | 1448.77 | 1465.81 | 1462.85 | 1399.96 | 1330.69 | 1296.04 | 1268.73 | 1216.14 | 1184.03 | -264.74 | -0.18 |
| *Strong opioids* | 1109.92 | 1116.26 | 1112.18 | 1057.53 | 991.50 | 956.22 | 922.50 | 877.72 | 844.76 | -265.16 | -0.24 |
| *Long-acting opioids* | 627.61 | 646.37 | 652.13 | 625.24 | 593.68 | 573.53 | 545.31 | 509.73 | 488.99 | -138.63 | -0.22 |
| *Morphine* | 241.41 | 256.95 | 265.72 | 260.03 | 246.99 | 242.64 | 231.67 | 218.59 | 210.78 | -30.64 | -0.13 |
| *Hydromorphone* | 0.92 | 0.85 | 0.83 | 0.74 | 0.68 | 0.61 | 0.60 | 0.47 | 0.47 | -0.45 | -0.49 |
| *Oxycodone* | 113.43 | 118.35 | 122.66 | 122.10 | 120.23 | 116.83 | 112.07 | 105.10 | 107.92 | -5.51 | -0.05 |
| *Dihydrocodeine* | 51.58 | 50.04 | 47.34 | 43.62 | 41.34 | 39.07 | 38.79 | 35.79 | 34.33 | -17.25 | -0.33 |
| *Pethidine* | 0.52 | 0.49 | 0.46 | 0.38 | 0.35 | 0.28 | 0.17 | 0.11 | 0.10 | -0.42 | -0.80 |
| *Fentanyl* | 176.80 | 179.57 | 177.67 | 166.07 | 153.23 | 140.85 | 129.06 | 116.23 | 105.93 | -70.87 | -0.40 |
| *Buprenorphine* | 68.48 | 72.47 | 76.82 | 77.56 | 77.86 | 79.92 | 79.44 | 79.30 | 76.83 | 8.34 | 0.12 |
| *Tramadol* | 502.49 | 478.80 | 456.38 | 416.84 | 376.98 | 359.09 | 352.51 | 341.41 | 327.61 | -174.88 | -0.35 |
| *Meptazinol* | 1.71 | 1.64 | 1.53 | 1.39 | 1.26 | 1.16 | 1.01 | 0.81 | 0.75 | -0.96 | -0.56 |
| *Tapentadol* | 5.86 | 8.76 | 11.65 | 13.80 | 15.18 | 16.01 | 16.99 | 16.50 | 15.13 | 9.26 | 1.58 |
| *Codeine* | 285.58 | 297.87 | 301.81 | 297.42 | 296.59 | 299.59 | 306.43 | 301.82 | 304.19 | 18.62 | 0.07 |

**Proportional share of analgesic prescribing**

|  | **2014** | **2015** | **2016** | **2017** | **2018** | **2019** | **2020** | **2021** | **2022** |
| --- | --- | --- | --- | --- | --- | --- | --- | --- | --- |
| Opioids | 0.40 | 0.40 | 0.39 | 0.40 | 0.40 | 0.41 | 0.41 | 0.41 | 0.40 |
| *Strong opioids* | 0.16 | 0.16 | 0.16 | 0.16 | 0.16 | 0.16 | 0.17 | 0.16 | 0.16 |
| *Long-acting opioids* | 0.07 | 0.07 | 0.08 | 0.08 | 0.08 | 0.08 | 0.08 | 0.08 | 0.08 |
| *Morphine* | 0.04 | 0.04 | 0.04 | 0.05 | 0.05 | 0.05 | 0.05 | 0.05 | 0.05 |
| *Hydromorphone* | <0.01 | <0.01 | <0.01 | <0.01 | <0.01 | <0.01 | <0.01 | <0.01 | <0.01 |
| *Oxycodone* | 0.01 | 0.01 | 0.01 | 0.02 | 0.02 | 0.02 | 0.02 | 0.02 | 0.02 |
| *Dihydrocodeine* | 0.04 | 0.04 | 0.04 | 0.04 | 0.03 | 0.03 | 0.03 | 0.03 | 0.03 |
| *Pethidine* | <0.01 | <0.01 | <0.01 | <0.01 | <0.01 | <0.01 | <0.01 | <0.01 | <0.01 |
| *Fentanyl* | 0.01 | 0.01 | 0.01 | 0.01 | 0.01 | 0.01 | 0.01 | 0.01 | 0.01 |
| *Buprenorphine* | 0.02 | 0.02 | 0.02 | 0.02 | 0.02 | 0.02 | 0.02 | 0.02 | 0.02 |
| *Tramadol* | 0.08 | 0.07 | 0.07 | 0.07 | 0.06 | 0.06 | 0.06 | 0.06 | 0.06 |
| *Meptazinol* | <0.01 | <0.01 | <0.01 | <0.01 | <0.01 | <0.01 | <0.01 | <0.01 | <0.01 |
| *Tapentadol* | <0.01 | <0.01 | <0.01 | <0.01 | <0.01 | <0.01 | <0.01 | <0.01 | <0.01 |
| *Codeine* | 0.20 | 0.20 | 0.20 | 0.20 | 0.20 | 0.21 | 0.21 | 0.21 | 0.21 |
| Systemic NSAIDs | 0.13 | 0.13 | 0.12 | 0.11 | 0.10 | 0.10 | 0.09 | 0.10 | 0.10 |
| *Non-selective NSAIDs* | 0.13 | 0.12 | 0.11 | 0.10 | 0.10 | 0.09 | 0.08 | 0.09 | 0.09 |
| *Coxib* | 0.01 | 0.01 | 0.01 | 0.01 | 0.01 | 0.01 | 0.01 | 0.01 | 0.01 |
| Paracetamol (incl. combinations) | 0.43 | 0.41 | 0.40 | 0.38 | 0.36 | 0.35 | 0.36 | 0.34 | 0.33 |
| *Paracetamol (excl. combinations)* | 0.24 | 0.23 | 0.22 | 0.20 | 0.19 | 0.18 | 0.18 | 0.17 | 0.16 |
| Gabapentin | 0.05 | 0.06 | 0.06 | 0.07 | 0.08 | 0.08 | 0.08 | 0.08 | 0.08 |
| Pregabalin | 0.04 | 0.05 | 0.05 | 0.06 | 0.07 | 0.07 | 0.08 | 0.09 | 0.08 |
| Amitriptyline | 0.06 | 0.06 | 0.06 | 0.07 | 0.07 | 0.08 | 0.08 | 0.09 | 0.10 |
| Topical analgesics | 0.06 | 0.06 | 0.07 | 0.07 | 0.07 | 0.06 | 0.05 | 0.05 | 0.05 |
| *Topical NSAIDs* | 0.06 | 0.06 | 0.06 | 0.06 | 0.06 | 0.06 | 0.04 | 0.04 | 0.05 |
| *Lidocaine* | <0.01 | <0.01 | <0.01 | <0.01 | <0.01 | <0.01 | <0.01 | <0.01 | <0.01 |
| *Capsaicin* | <0.01 | <0.01 | <0.01 | <0.01 | <0.01 | <0.01 | <0.01 | <0.01 | <0.01 |
| Antimigraines | 0.03 | 0.02 | 0.02 | 0.02 | 0.02 | 0.03 | 0.03 | 0.03 | 0.03 |

**Proportional share of opioid prescribing**

|  | **2014** | **2015** | **2016** | **2017** | **2018** | **2019** | **2020** | **2021** | **2022** |
| --- | --- | --- | --- | --- | --- | --- | --- | --- | --- |
| Strong opioids | 0.40 | 0.40 | 0.41 | 0.41 | 0.40 | 0.40 | 0.40 | 0.40 | 0.40 |
| Long-acting opioids | 0.18 | 0.19 | 0.19 | 0.20 | 0.20 | 0.20 | 0.20 | 0.20 | 0.20 |
| Morphine | 0.09 | 0.10 | 0.11 | 0.12 | 0.12 | 0.12 | 0.12 | 0.13 | 0.13 |
| Hydromorphone | <0.01 | <0.01 | <0.01 | <0.01 | <0.01 | <0.01 | <0.01 | <0.01 | <0.01 |
| Oxycodone | 0.03 | 0.03 | 0.04 | 0.04 | 0.04 | 0.04 | 0.05 | 0.05 | 0.05 |
| Dihydrocodeine | 0.11 | 0.10 | 0.09 | 0.09 | 0.09 | 0.08 | 0.08 | 0.07 | 0.07 |
| Pethidine | <0.01 | <0.01 | <0.01 | <0.01 | <0.01 | <0.01 | <0.01 | <0.01 | <0.01 |
| Fentanyl | 0.03 | 0.03 | 0.03 | 0.03 | 0.03 | 0.02 | 0.02 | 0.02 | 0.02 |
| Buprenorphine | 0.04 | 0.04 | 0.05 | 0.05 | 0.05 | 0.05 | 0.05 | 0.05 | 0.05 |
| Tramadol | 0.20 | 0.19 | 0.18 | 0.17 | 0.16 | 0.16 | 0.16 | 0.16 | 0.15 |
| Meptazinol | <0.01 | <0.01 | <0.01 | <0.01 | <0.01 | <0.01 | <0.01 | <0.01 | <0.01 |
| Tapentadol | <0.01 | <0.01 | <0.01 | <0.01 | <0.01 | <0.01 | <0.01 | <0.01 | <0.01 |
| Codeine | 0.49 | 0.50 | 0.50 | 0.50 | 0.51 | 0.51 | 0.52 | 0.52 | 0.53 |
